# Innate lymphoid cell composition associates with COVID-19 disease severity

**DOI:** 10.1101/2020.10.13.20211367

**Authors:** Marina García, Efthymia Kokkinou, Anna Carrasco García, Tiphaine Parrot, Laura M. Palma Medina, Kimia T. Maleki, Wanda Christ, Renata Varnaitė, Iva Filipovic, Hans-Gustaf Ljunggren, Niklas K. Björkström, Elin Folkesson, Olav Rooyackers, Lars I. Eriksson, Anders Sönnerborg, Soo Aleman, Kristoffer Strålin, Sara Gredmark-Russ, Jonas Klingström, Jenny Mjösberg, the Karolinska KI/K COVID-19 Study Group

## Abstract

**Objectives:** The role of innate lymphoid cells (ILCs) in coronavirus disease 2019 (COVID-19), caused by severe acute respiratory syndrome coronavirus 2 (SARS-CoV-2), is unknown. Understanding the immune response in COVID-19 could contribute to unravel the pathogenesis and identification of treatment targets. To describe the phenotypic landscape of circulating ILCs in COVID-19 patients and to identify ILC phenotypes correlated to serum biomarkers, clinical markers, and laboratory parameters relevant in COVID-19.

**Methods:** Blood samples collected from moderately (n=11) and severely ill (n=12) COVID-19 patients as well as healthy control donors (n=16), were analyzed with 18-parameter flow cytometry. Using supervised and unsupervised approaches, we examined the ILC activation status and homing profile. Clinical and laboratory parameters were obtained from all COVID-19 patients and serum biomarkers were analyzed with multiplex immunoassays.

**Results:** ILCs were largely depleted from the circulation of COVID-19 patients compared with healthy controls. Remaining circulating ILCs from patients revealed increased frequencies of ILC2 in moderate COVID-19, with a concomitant decrease of ILC precursors (ILCp), as compared with controls. ILC2 and ILCp showed an activated phenotype with increased CD69 expression, whereas expression levels of the chemokine receptors CXCR3 and CCR4 were significantly altered in ILC2 and ILCp, and ILC1, respectively. The activated ILC profile of COVID-19 patients was associated with soluble inflammatory markers, while frequencies of ILC subsets were correlated with laboratory parameters that reflect the disease severity.

**Conclusion:** This study provides insights into the potential role of ILCs in immune responses against SARS-CoV-2, particularly linked to the severity of COVID-19.

## INTRODUCTION

Coronavirus disease 2019 (COVID-19), caused by the novel severe acute respiratory syndrome coronavirus 2 (SARS-CoV-2), is of global concern and major efforts to identify effective treatments and vaccines are currently underway^1–4^. SARS-CoV-2 affects mainly the respiratory tract and infected individuals often present with flu-like symptoms. While some patients recover within weeks, others progress to a severe stage of disease, typically including severe respiratory distress and hypoxia with later multi-organ failure, occasionally leading to death^5,6^. Notably, a gradual progression to more severe disease stages coincides with the development of a hyperactivated immune response, triggering a systemic cytokine storm circulatory impairment, sometimes leading to circulatory shock^7–9^. Thus, there are reasons to believe that immunopathology involving the innate immune system plays a potent role behind severe morbidity and mortality in the critically ill COVID-19 patients. Currently, little information is available on the role of innate immune cells in COVID-19. Therefore, here we studied the role of innate lymphoid cell (ILC) in hospitalized COVID-19 patients.

ILCs are innate lymphocytes which, unlike T- and B-cells, lack rearranged antigen specific receptors and cell surface markers associated with other lymphoid and myeloid lineages^10^. ILCs are categorized into five major groups: natural killer (NK) cells, ILC1, ILC2, ILC3 and Lymphoid Tissue Inducer (LTi) cells^10^. Based on their transcription factor dependence and functional characteristics, ILC1^11,12^, ILC2^13–15^ and ILC3^16^ are considered the innate counterparts of the specialized subsets of CD4^+^ T cells, i.e. Th1, Th2 and Th17 cells, respectively, while NK cells mirror the cytotoxic functions of CD8^+^ T cells^10^. Although ILCs exert their function primarily in tissues, distinct subsets of ILCs are found circulating in blood^17,18^. The major blood ILC population is the precursor ILC (ILCp), with the capacity to home to peripheral tissues and differentiate to mature ILC subsets^18^. However, human blood also harbors the committed ILC lineages ILC1 and ILC2. ILC2 in blood express CRTH2 and CD161 and are dependent on the GATA binding protein-3 (GATA-3) transcription factor^19^. IFN-γ-producing ILCs, reminiscent of ILC1, have been identified in peripheral blood^20^. While their characterization has been challenging due to the lack of unique surface markers^21^, studies have shown that blood ILC1 that produce IFN-γ express the chemokine receptor CXCR3^20^. Hence, CXCR3 serves as a marker to define the ILC1 subset in peripheral blood.

While little is known regarding ILCs in the context of human respiratory viral infections, studies performed in mice highlight their contribution during acute viral infection. More specifically, ILC2 have been suggested to accumulate in the lung of influenza virus-infected mice and promote lung homeostasis and tissue repair^22^. In contrast, ILC2 in concordance with T cells, were reported to contribute to allergic airway inflammation induced by influenza virus in mice^23^. In humans, ILCs have been investigated in acutely HIV-1-infected individuals where they were found to be depleted from the circulation^24^. To the best of our knowledge, ILCs have not yet been investigated in a human respiratory viral disease.

Understanding the immunopathogenesis of COVID-19 is urgently needed in order to help tackle the current pandemic. In the present study, we report a profound reduction in number of ILCs within the systemic circulation of COVID-19 patients. The remaining ILCs show dysregulated expression of markers associated with activation and migration. Furthermore, ILC frequencies are correlated to clinical parameters related to COVID-19 disease severity.

## MATERIAL AND METHODS

### Study participants and sampling

As a part of the Karolinska KI/K COVID-19 Immune Atlas, 23 COVID-19 patients (6 females and 17 males; median age 57 years; age range 18 - 74 years) positive for SARS-CoV-2 RNA by diagnostic RT-qPCR and hospitalized at the Karolinska University Hospital (Stockholm, Sweden) were included in the present study. Patients were classified as COVID-19 moderate (n=11) and severe (n=12), based on the peak supplementary oxygen received during hospitalization until the time of inclusion in the study. The moderate group required low or no supplementary oxygen (low oxygen flow, ≤15L/min), out of whom 8 were hospitalized in the regular ward and 3 in the intensive care unit (ICU). These patients had median ordinal scale of 5 (IQR 4-5) at sampling. The severe group consisted of patients that were hospitalized in the ICU and required mechanical ventilation or extracorporeal membrane oxygenation (ECMO; one patient) provided within an intensive care setting. These patients had a median ordinal scale score of 7 (IQR 7-7) at sampling. This classification was in line with the 8-category ordinal scale described by Beigel *et al* as well as in the WHO guidelines^25,26^.

Peripheral blood sample from all COVID-19 patients was collected 5 to 24 days (median 14 days; IQR 5-24 days) after symptom debut and 0 to 8 days after hospitalization.

Sixteen donors (6 females and 10 males; median age 54 years; age range between 34 - 69 years), all SARS-CoV-2 IgG seronegative and symptom-free at the day of sampling, were included as a control group for this study. Detailed donor characteristics are summarized in Tables 1-2.

**Table 1.**
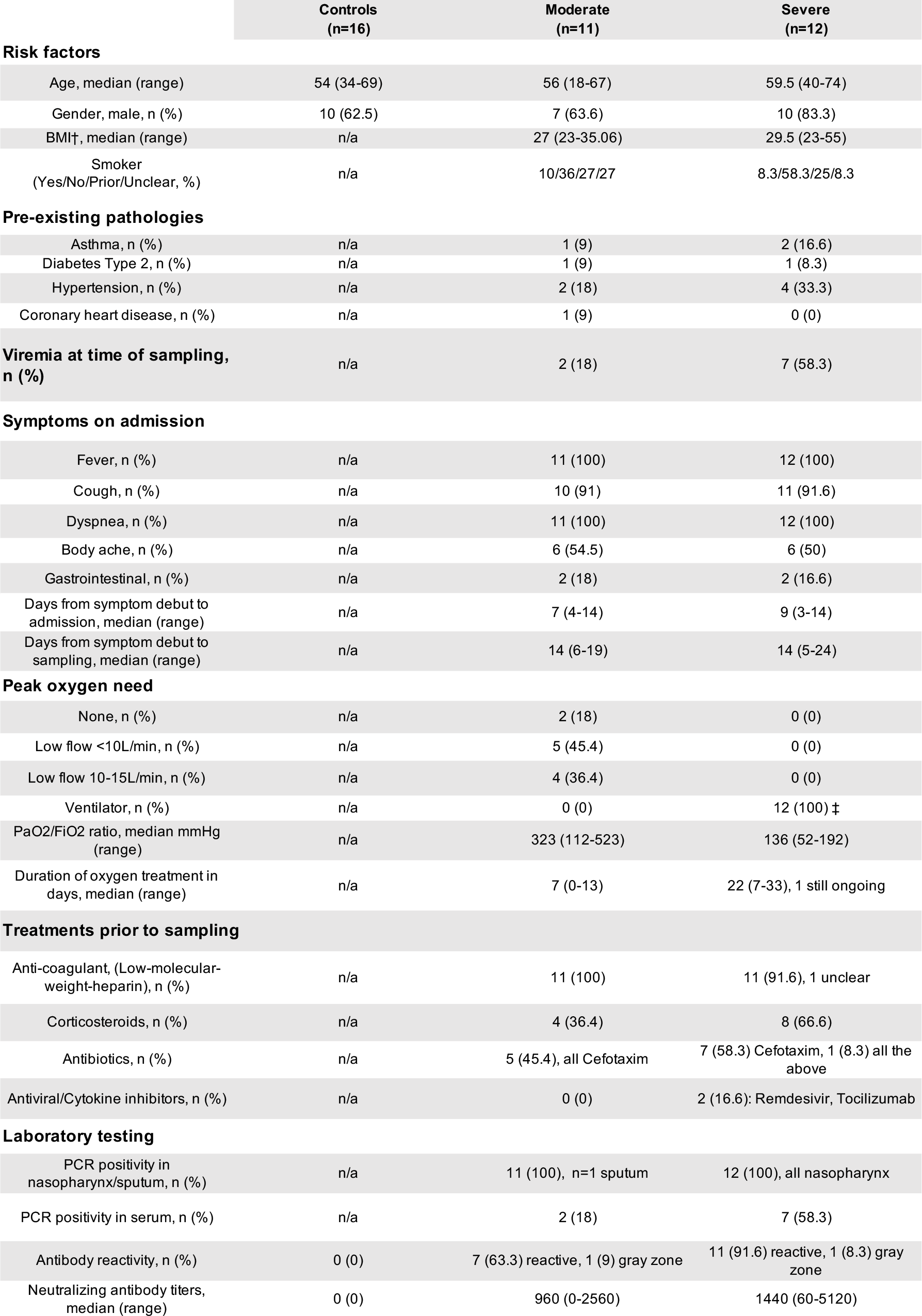
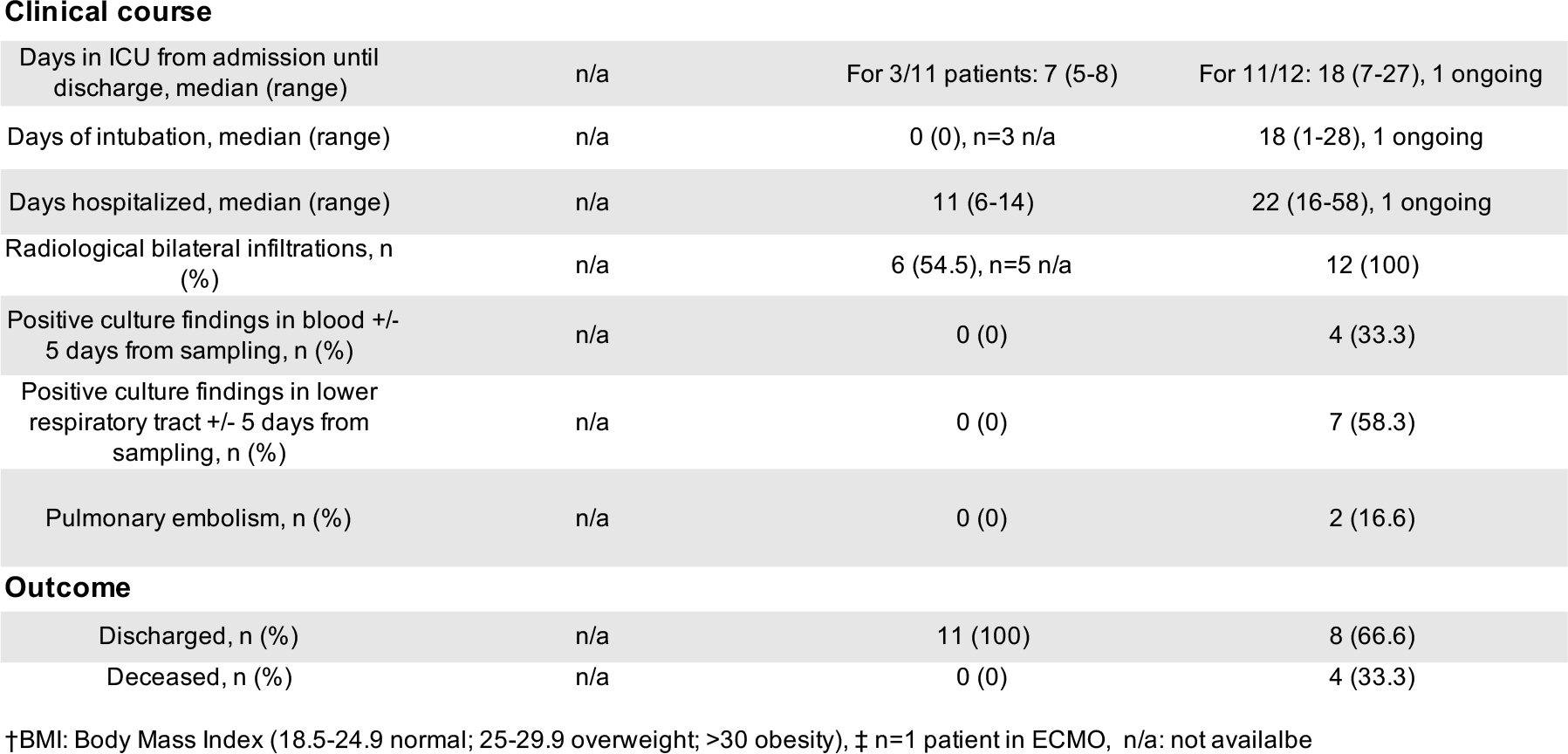
Donor characteristics

|  | Controls<br>(n=16) | Moderate<br>(n=11) | Severe<br>(n=12) |
| --- | --- | --- | --- |
| **Risk factors** |  |  |  |
| Age, median (range) | 54 (34-69) | 56 (18-67) | 59.5 (40-74) |
| Gender, male, n (%) | 10 (62.5) | 7 (63.6) | 10 (83.3) |
| BMI†, median (range) | n/a | 27 (23-35.06) | 29.5 (23-55) |
| Smoker<br>(Yes/No/Prior/Unclear, %) | n/a | 10/36/27/27 | 8.3/58.3/25/8.3 |
| **Pre-existing pathologies** |  |  |  |
| Asthma, n (%) | n/a | 1 (9) | 2 (16.6) |
| Diabetes Type 2, n (%) | n/a | 1 (9) | 1 (8.3) |
| Hypertension, n (%) | n/a | 2 (18) | 4 (33.3) |
| Coronary heart disease, n (%) | n/a | 1 (9) | 0 (0) |
| **Viremia at time of sampling,**<br>**n (%)** | n/a | 2 (18) | 7 (58.3) |
| **Symptoms on admission** |  |  |  |
| Fever, n (%) | n/a | 11 (100) | 12 (100) |
| Cough, n (%) | n/a | 10 (91) | 11 (91.6) |
| Dyspnea, n (%) | n/a | 11 (100) | 12 (100) |
| Body ache, n (%) | n/a | 6 (54.5) | 6 (50) |
| Gastrointestinal, n (%) | n/a | 2 (18) | 2 (16.6) |
| Days from symptom debut to<br>admission, median (range) | n/a | 7 (4-14) | 9 (3-14) |
| Days from symptom debut to<br>sampling, median (range) | n/a | 14 (6-19) | 14 (5-24) |
| **Peak oxygen need** |  |  |  |
| None, n (%) | n/a | 2 (18) | 0 (0) |
| Low flow <10L/min, n (%) | n/a | 5 (45.4) | 0 (0) |
| Low flow 10-15L/min, n (%) | n/a | 4 (36.4) | 0 (0) |
| Ventilator, n (%) | n/a | 0 (0) | 12 (100) ‡ |
| PaO2/FiO2 ratio, median mmHg<br>(range) | n/a | 323 (112-523) | 136 (52-192) |
| Duration of oxygen treatment in<br>days, median (range) | n/a | 7 (0-13) | 22 (7-33), 1 still ongoing |
| **Treatments prior to sampling** |  |  |  |
| Anti-coagulant, (Low-molecular-<br>weight-heparin), n (%) | n/a | 11 (100) | 11 (91.6), 1 unclear |
| Corticosteroids, n (%) | n/a | 4 (36.4) | 8 (66.6) |
| Antibiotics, n (%) | n/a | 5 (45.4), all Cefotaxim | 7 (58.3) Cefotaxim, 1 (8.3) all the<br>above |
| Antiviral/Cytokine inhibitors, n (%) | n/a | 0 (0) | 2 (16.6): Remdesivir, Tocilizumab |
| **Laboratory testing** |  |  |  |
| PCR positivity in<br>nasopharynx/sputum, n (%) | n/a | 11 (100), n=1 sputum | 12 (100), all nasopharynx |
| PCR positivity in serum, n (%) | n/a | 2 (18) | 7 (58.3) |
| Antibody reactivity, n (%) | 0 (0) | 7 (63.3) reactive, 1 (9) gray zone | 11 (91.6) reactive, 1 (8.3) gray<br>zone |
| Neutralizing antibody titers,<br>median (range) | 0 (0) | 960 (0-2560) | 1440 (60-5120) |
| Clinical course |  |  |  |
| Days in ICU from admission until discharge, median (range) | n/a | For 3/11 patients: 7 (5-8) | For 11/12: 18 (7-27), 1 ongoing |
| Days of intubation, median (range) | n/a | 0 (0), n=3 n/a | 18 (1-28), 1 ongoing |
| Days hospitalized, median (range) | n/a | 11 (6-14) | 22 (16-58), 1 ongoing |
| Radiological bilateral infiltrations, n (%) | n/a | 6 (54.5), n=5 n/a | 12 (100) |
| Positive culture findings in blood +/- 5 days from sampling, n (%) | n/a | 0 (0) | 4 (33.3) |
| Positive culture findings in lower respiratory tract +/- 5 days from sampling, n (%) | n/a | 0 (0) | 7 (58.3) |
| Pulmonary embolism, n (%) | n/a | 0 (0) | 2 (16.6) |
| Outcome |  |  |  |
| Discharged, n (%) | n/a | 11 (100) | 8 (66.6) |
| Deceased, n (%) | n/a | 0 (0) | 4 (33.3) |

†BMI: Body Mass Index (18.5-24.9 normal; 25-29.9 overweight; >30 obesity), ‡ n=1 patient in ECMO, n/a: not available

**Table 2.**
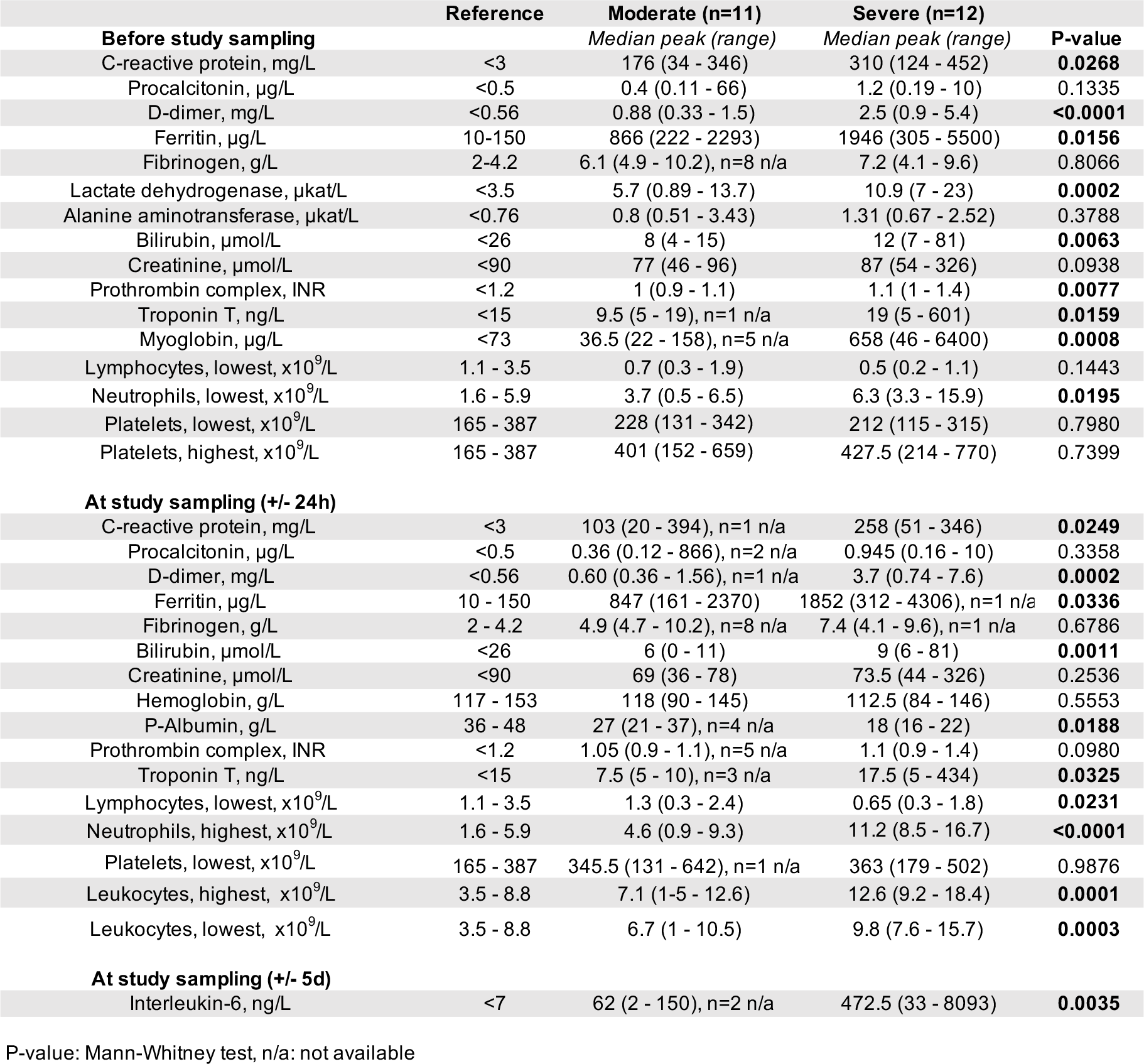
Laboratory parameters of COVID-19 patients

This study was approved by the Swedish Ethical Review Authority and conducted according to the Declaration of Helsinki. All patients or next of kin and control donors provided oral and/or written informed consent in line with the ethical approval.

### Serum collection and cell isolation from whole blood

Peripheral blood mononuclear cells (PBMCs) from control donors and COVID-19 patients were isolated from heparinized anti-coagulated blood in SepMate tubes (Stemcell Technologies) using gradient density centrifugation, according to manufacturer’s instructions. Briefly, 15ml of Lymphoprep™ (Stemcell Technologies) was pipetted below the SepMate insert and 20ml of diluted whole blood was dispensed on top. Tubes were centrifuged at 1200 *g* for 10 min. The supernatant including PBMCs was carefully decanted into a new tube, followed by two washes with PBS containing 10% FCS. Platelets were removed by centrifuging the samples at 400 *g* for 10 min. Pellets were resuspended, cells counted and subsequently stained for flow cytometry.

Serum was collected from COVID-19 patients and control donors in BD Vacutainer serum tubes with spray-coated silica (BD Biosciences). After coagulation for up to 2 hours at room temperature (RT), serum was isolated by centrifugation at 2000 *g* for 10 min and immediately stored at −80 °C for later analysis.

### Cell staining and flow cytometry analysis

Freshly isolated PBMCs were stained with dead cell marker (Invitrogen) and fluorochrome-conjugated antibodies directed against intracellular and surface markers (See Table S1 in the Online Repository) for 20 min at RT in the dark. Cells were washed two times with 150 ul of PBS with 10% FCS. Next, pellets were resuspended in 100 ul of BD FACS Lysing Solution (BD Biosciences) and incubated for 10 min at RT, for cell fixation. After washing with PBS with 10% FCS, cells were permeabilized using 100 ul of BD Perm2 Permeabilizing Solution (BD Biosciences) for 10 min at RT. Subsequently, antibodies were added for intracellular staining and incubated for 30 min at RT in the dark. Cells were then washed with 150 ul of PBS with 10% FCS and incubated in a 1% formaldehyde solution for 2h, washed and resuspended in PBS containing 10% FCS.

The antibodies and dead cell marker used for flow cytometry staining are listed in Supplementary Table III. Samples were acquired on a BD LSR Fortessa™. Flow cytometric analysis was performed using FlowJo version 10.6.2 (TreeStar).

### UMAP analysis

To ensure unbiased manual gating, a blinded analysis was implemented, whereby all FCS3.0 files were renamed and coded by one person and blindly analyzed by another person. All samples were compensated electronically and gatings were based on fluorescent-minus-one (FMO) or negative controls. After all gatings were performed, samples were decoded and statistical analysis between groups and unsupervised analysis was performed. For unsupervised analysis, the following FlowJo plugins were used: DownSample (v.1.1), UMAP (v.2.2), Phenograph (v.2.4) and ClusterExplorer (v.1.2.2) (all FlowJo LLC). First, 133 events per patients were downsampled from the total ILC gate (Fig. 2SB). The new generated FCS files were labelled according to control or patient group (moderate or severe COVID-19) and concatenated per group. Subsequently, all groups were taken to the same number of events, by downsampling to the number of events present in the group with the least number of events. The three new FCS files corresponding to control, moderate and severe COVID-19 patients were then concatenated for dimensionality reduction analysis using UMAP. UMAP was run including the markers CRTH2, CD161, CD117, CD69, HLA-DR, Ki-67, CD45RA, CD62L, CCR4, CCR6, CXCR3, CD56, NKp44 and using the following parameters: metric = euclidean, nearest neighbors = 15, and minimum distance = 0.5. Clusters of phenotypically related cells were identified using Phenograph plugin and including the same markers as for UMAP and parameters k = 30 and Run ID = auto. Finally, ClusterExplorer plugin was used to study the phenotype of the different clusters and to generate heatmaps of marker expression from those clusters.

**Figure 1.**
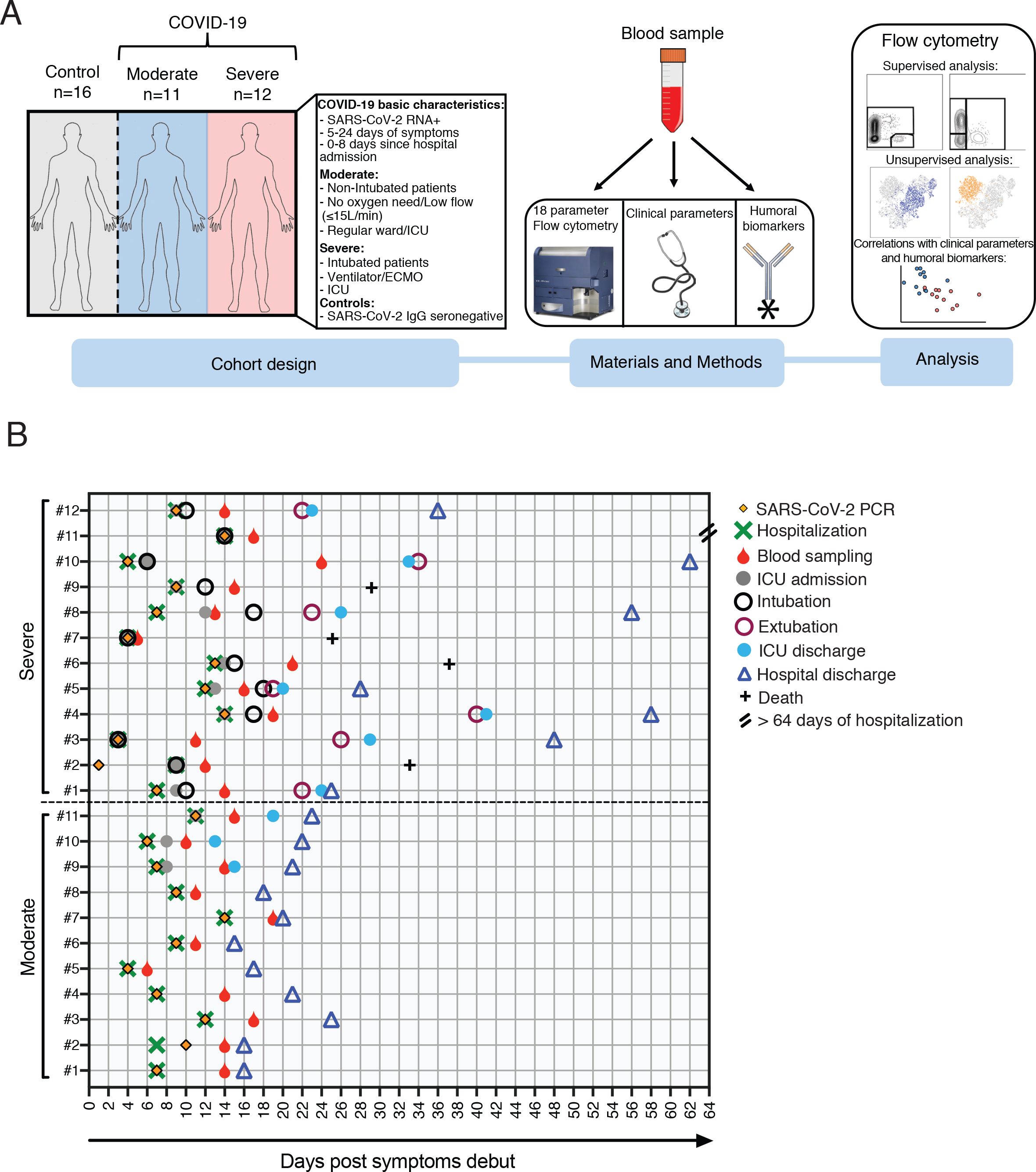
Experimental design and COVID-19 cohort characteristics. **(A)** Schematic representation of the cohort characteristics (left), materials and methods (middle) and the type of analysis performed using the flow cytometric data (right). **(B)** Graphical overview of all COVID-19 patients (n=23) illustrating clinical events from the day of the patient symptom debut. Depicted are the day of the SARS-CoV-2 PCR test, hospitalization and blood sampling. For all the severe patients and three of the moderate patients the day of the intensive care unit (ICU) admission and discharge is indicated. Furthermore, the day of intubation/extubation of the severe patients is shown. Lastly, depicted is the day of hospital discharge for 22 out of 23 patients and the number of deceased patients (n=4). One patient from the severe group (#11) is still in ECMO with ongoing hospitalization (>64 days).

**Figure 2.**
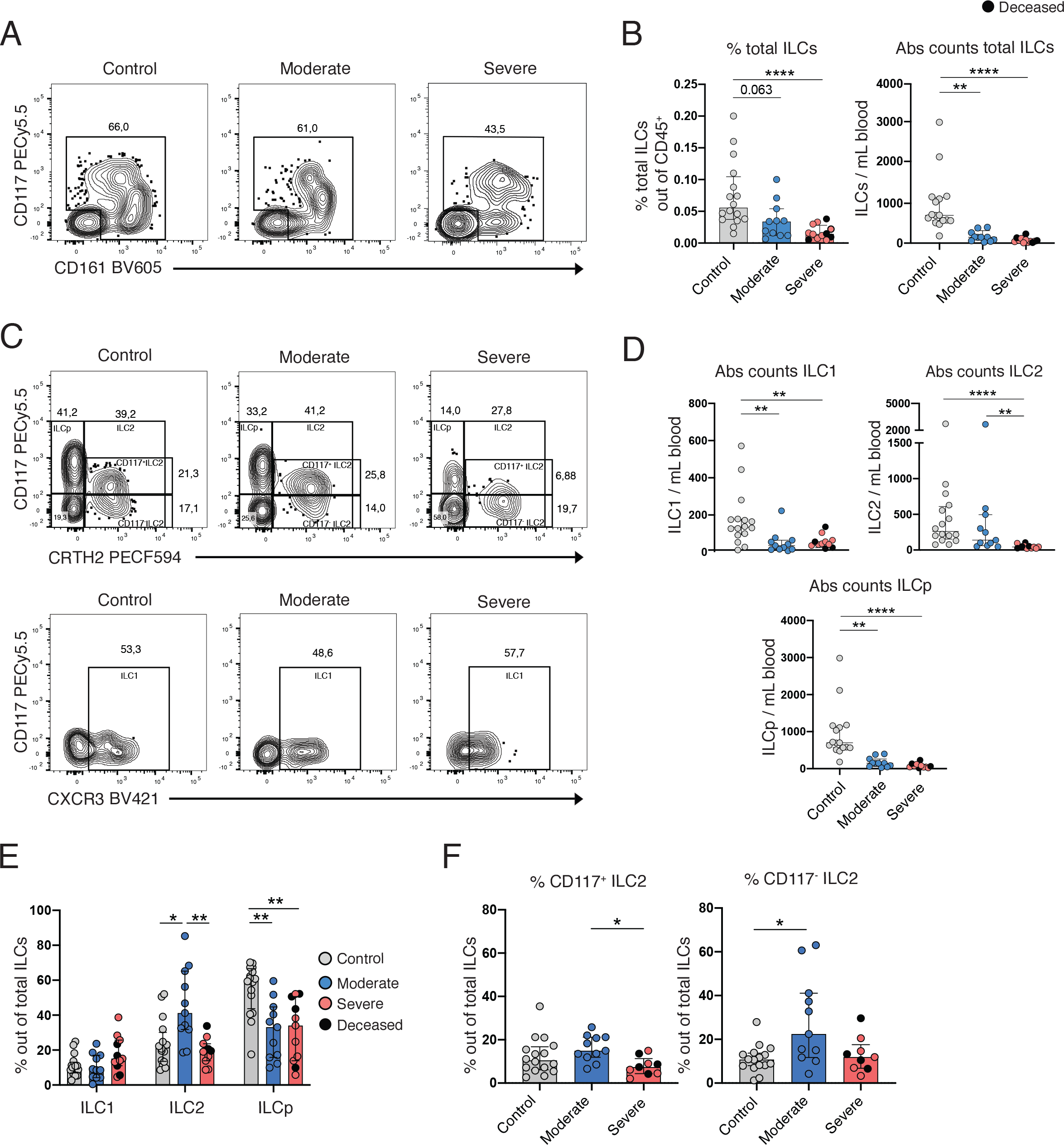
Depletion and altered frequency of ILCs in the peripheral blood of COVID-19 patients. **(A)** Representative flow cytometry plots depicting the gate for the identification of the total ILCs in one control donor, one moderate and one severe COVID-19 patient. (**B)** Bar plot summaries of (A) the percentage (left) and absolute counts (per mL blood) (right) of total ILCs in control donors (n=16), moderate (n=11) and severe (n=12) COVID-19 patients. **(C)** Representative flow cytometry plots depicting total ILCs gated as: ILCp, ILC2 (CD117^+^/^-^) (upper row) and ILC1 (lower row) in one control donor, one moderate and one severe COVID-19 patient. **(D)** Bar plot summaries of (C) absolute counts of ILC1, ILC2 and ILCp subsets (per mL blood) in control donors (n=16), moderate (n=11) and severe (n=11) COVID-19 patients. **(E)** Bar plot summaries of (C) the percentage of the ILC1, ILC2 and ILCp of total ILCs in control donors (n=16), moderate (n=11) and severe (n=11) COVID-19 patients. **(F)** Bar plot summaries of (C) the percentage of CD117^+^ and CD117^-^ ILC2 of total ILCs in control donors (n=16), moderate (n=11) and severe (n=9) COVID-19 patients. **(B, C, E, F)** statistical differences were tested using Kruskal-Wallis test followed by Dunn’s multiple comparisons. Numbers in flow cytometry plots indicate percentage of cells within the mother gate. Bar graphs are shown as median ± IQR, *p < 0.05, ** p < 0.01, *** p < 0.001. Patients with low cell numbers (less than 20 events) in the corresponding gate were removed from the analysis.

### Absolute cell counts

Absolute numbers of ILCs in peripheral blood were obtained using BD Multitest™ 6-color TBNK reagents with bead-containing BD Trucount™ tubes (BD Biosciences), according to manufacturer’s instructions. Briefly, 50 µL of anti-coagulated whole blood were added into Trucount tubes within 3h after blood extraction. Then, an antibody mix was added for staining. After 15 minutes of incubation at RT in the dark, stained whole blood was fixed and red blood cells lysed with 1X BD FACS Lysing Solution (BD Biosciences) for 2h at RT. Samples were then fixed with 1% PFA for 2h prior to acquisition in a FACSymphony A5 (BD Biosciences) flow cytometer. Absolute ILC cell counts were calculated using the formula:

ILC (#/mL) = [(# lymphocytes events acquired * total #beads in tube * 1000) / (#beads acquired * volume of whole blood stained (µL))] * % ILC out of lymphocytes. ILC1, ILC2 and ILCp absolute counts were calculated using their frequencies relative to total ILCs.

### Analysis of serum biomarkers

Heat-inactivated (56°C for 30 min) serum from all patients and controls were analyzed for soluble protein biomarkers using proximity extension assay (PEA) technology (Olink AB, Uppsala, Sweden). The samples were analyzed using the inflammation (v.3022) panel, including a total of 92 analytes. Data are expressed as normalized protein expression (NPX) values. NPX is an arbitrary unit on a Log2 scale to normalize data to minimize both intra-assay and inter-assay variation.

Additionally, several soluble analytes were also measured in serum or plasma by use of custom made multiplex Magnetic Luminex Screening assays (R&D Systems), according to the manufacturer’s instructions. Serum and plasma were diluted 1:2 prior to analysis in multiplex.

Analytes from Olink data and Magnetic Luminex Screening assays that had more than 33% and 25% of missing values, respectively, were excluded from analysis. Left-censored data from the multiplex analysis were imputed using *GSimp* package^27^ in R (v. 3.6.0)^28^.

### Clinical parameters and serology

Serum samples from all patients and controls were analyzed for presence of SARS-CoV-2 antibodies, as recently described^29^.

Micro-neutralization assay for measurement of neutralizing antibody titers was performed as previously described^30^. Briefly, heat inactivated serum (56°C for 30 minutes) was diluted two-fold starting at 1:10, mixed with an equal volume of 200 TCID50 SARS-CoV-2 (50 µl diluted serum plus 50 µl virus) and incubated for 1 hour at 37 °C and 5% CO2. Mixtures were then added to Vero E6 cells and incubated at 37 °C 5% CO2 for four days. Cells were inspected for signs of cytopathic effect (CPE) by optical microscopy. Results were expressed as the arithmetic mean of the reciprocals of the highest neutralizing dilutions from the two duplicates for each sample.

### Statistical analysis

Statistical analyses were performed using Prism version 8.4.3 (GraphPad Software Inc.). For comparisons between three unpaired groups, Kruskal-Wallis test followed by Dunn’s multiple comparisons test was used. Correlation analyses were performed using Spearman’s rank correlation. Spearman’s correlation matrix was generated with R (v.4.0.2) using package corrplot (v.0.84). Statistical significance for differences between COVID-19 patients and healthy controls was determined by two-sided Mann-Whitney *U* test. p-values < 0.05 were considered statistically significant.

Principal component analysis (PCA) was performed in R (v.4.0.2; R Core Team, 2020) using packages Factoextra (v.1.0.7)^31^, FactoMineR (v.2.3)^32^, RColorBrewer (v.1.1-2)^33^, and ggplot2 (v.3.3.2)^34^. Data was normalized in R using the *scale* argument within the PCA function. Where data was missing, the values were imputed using package WaverR (v1.0)^35^ using 1000 repetitions.

## RESULTS

A total of twenty-three patients with either moderate (n=11) or severe (n=12) COVID-19 and sixteen control donors were included in the study (Fig. 1A). The COVID-19 patients showed profound perturbations in inflammatory markers, coagulation factors, organ/muscle damage markers as well as biochemical and hematological parameters (Fig. S1). A detailed summary of the patients’ clinical and laboratory parameters is presented in Tables 1-2. A timeline summarizing the major clinical events of the patients is shown in Fig. 1B.

For the identification and analysis of peripheral blood ILCs we used 18-parameter flow cytometry and a modification of a well-established gating strategy^36^ (Fig. S2A). We found that the relative frequencies and absolute counts of total CD127^+^ ILCs (hereafter referred to as total ILCs), as well as the absolute counts of the specific subsets ILC1, ILC2 and ILCp, were decreased in peripheral blood of COVID-19 patients as compared with controls (Fig. 2A-D). Further characterization of the remaining circulating ILC compartment showed reduced ILCp frequencies in COVID-19 patients, while the frequencies of ILC1 remained unchanged (Fig. 2C and E). Of note, the moderately ill group showed elevated frequencies of both total ILC2 (Fig. 2E) and CD117^-^ ILC2, as compared with the control group (Fig. 2F).

Taken together, we observed a general ILC depletion as well as compositional changes in the circulating ILC compartment of COVID-19 patients. Interestingly, changes in the relative frequency of ILCs differed between the moderate and severe group, prompting a more detailed phenotypical study of the ILCs.

To deepen our understanding of the differentiation, activation, and migration of ILCs during COVID-19, we assessed differentiation (CD56 and NKp44) (Fig. S3A and B) and activation (CD69, HLA-DR and Ki-67) markers as well as chemokine receptors (CCR4, CCR6 and CXCR3) and molecules associated with naivety and homing to lymphoid tissues (CD45RA and CD62L) on ILCs (Fig. 3A-H). Manual gating and unbiased principal component analysis (PCA) were used to simultaneously take into account the relative frequency of all the markers measured on ILCs.

**Figure 3.**
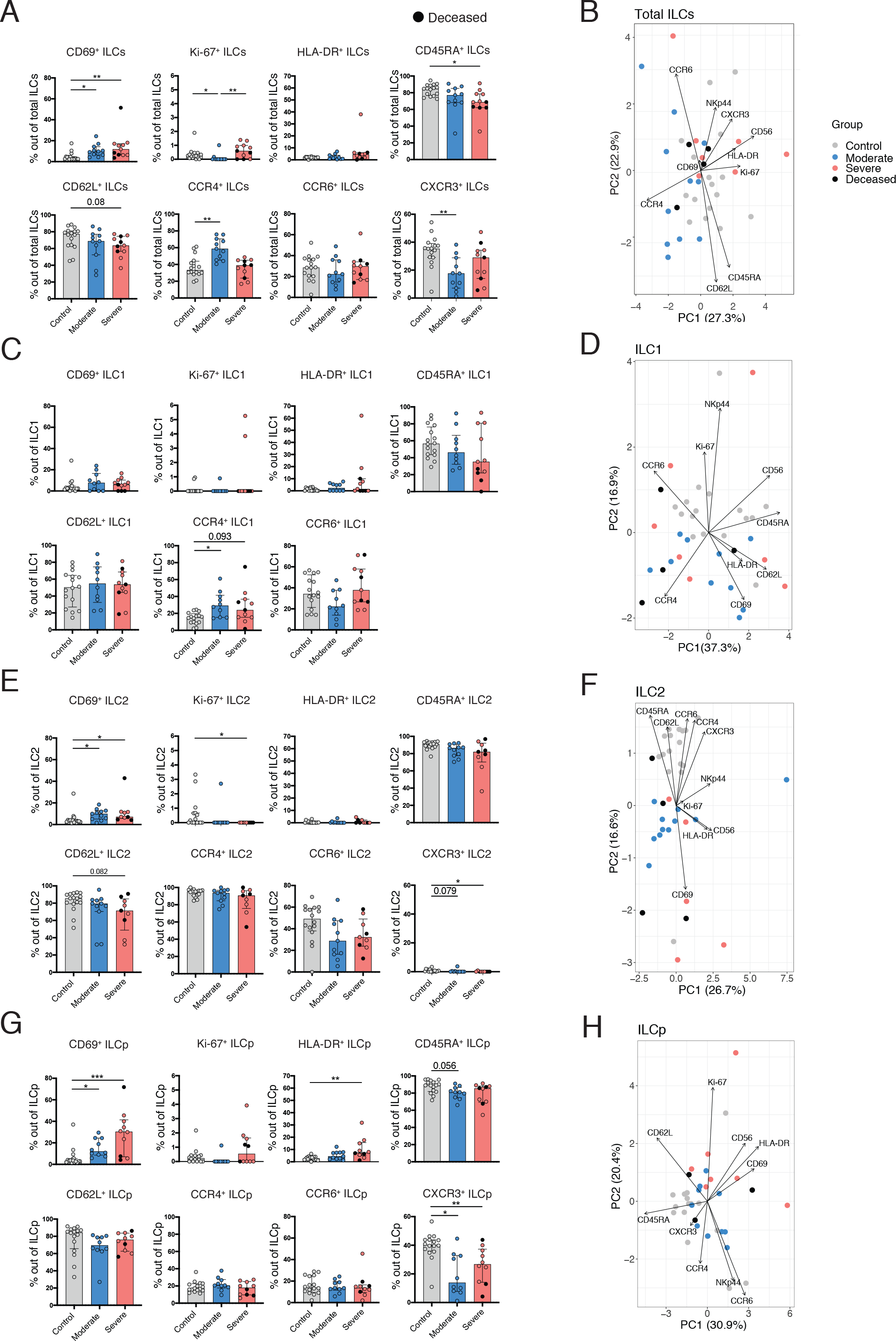
ILCs reveal an activated and migratory profile in peripheral blood of COVID-19 patients. **(A)** Bar plot summaries showing the percentages of the indicated markers in total ILCs in control donors (n=16), moderate (n=11) and severe (n=11) COVID-19 patients. **(B)** PCA plot of total ILCs from control donors (n=16), moderate (n=11) and severe (n=11) COVID-19 patients based on the cell surface markers presented in (A). **(C)** Bar plot summaries showing the percentages of the indicated markers in ILC1 subset in control donors (n=16), moderate (n=10) and severe (n=11) groups. **(D)** PCA plot of ILC1 showing the contribution of cell surface markers indicated in (C). **(E)** Bar plot summaries showing the percentages of the indicated markers in ILC2 subset in control donors (n=16), moderate (n=11) and severe (n=9) groups. **(F)** PCA plot of ILC2 showing the contribution of cell surface markers indicated in (E). **(G)** Bar plot summaries showing the percentages of the indicated markers in ILCp subset in control donors (n=16), moderate (n=10) and severe (n=10) groups. **(H)** PCA plot of ILCp showing the contribution of cell surface markers indicated in (G). **(A, C, E, G)**, statistical differences were tested using Kruskal-Wallis test followed by Dunn’s multiple comparisons. Bar graphs are shown as median ± IQR, *p < 0.05, ** p < 0.01, *** p < 0.001. Patients with low cell numbers (less than 20 events) in the corresponding gate were removed from the analysis. In PCA plots each dot represents one donor. Deceased patients in the severe group are indicated by a black dot.

This approach revealed a higher relative frequency of CD69-expressing total ILCs in patients as compared with controls (Fig. 3A). Additionally, Ki-67, a marker of cell proliferation, was increased in severe compared with moderate COVID-19 but decreased in moderate COVID-19 compared with controls (Fig. 3A). However, the overall Ki-67 expression level was low in all ILCs regardless of the study group, suggesting either a low proliferating capacity of ILCs in peripheral blood or migration of proliferative ILCs to tissues at an earlier stage. In line with an increase in CD69 in COVID-19 patients, we detected reduced frequencies of total ILCs expressing CD45RA and CD62L, two markers associated with ILC naivety^18^, in severe COVID-19 patients (Fig. 3A). Additionally, we observed alterations in chemokine receptor expression between controls and moderate COVID-19 patients, revealing a reduction in the percentage of CXCR3^+^ and an increase in CCR4^+^ ILCs. The latter likely reflects the increase in ILC2 frequencies in these patients (Fig. 2E), as CCR4 is particularly highly expressed on ILC2 (Fig. 3E). Despite these changes in ILC phenotypes, the PCA analysis could not discriminate between COVID-19 patients and controls on the basis of the total ILC data (Fig. 3B), suggesting that the COVID-19 related changes in total ILCs are ILC subset specific.

Indeed, we observed no changes in expression of markers associated with differentiation and activation in ILC1. There was, however, a slightly increased percentage of CCR4^+^ ILC1 cells in COVID-19 patients compared to controls (Fig. 3C). We also detected a reduction of CD56^+^ ILC1 in COVID-19 patients (Fig. S3A). PCA analysis illustrated these differences well, showing segregation of the ILC1 data on the basis of CCR4 for COVID-19 patients and CD56 for controls (Fig. 3D).

In contrast, ILC2 displayed an activated phenotype in COVID-19 patients, showing higher frequencies of CD69^+^ and a tendency towards reduced frequencies of CD62L^+^ cells, as compared with controls (Fig. 3E). Interestingly, Ki-67, generally expressed at very low levels, was decreased in relative frequency in ILC2 from severe COVID-19 patients compared to the control group, possibly reflecting tissue recruitment of such cells in severe COVID-19. Indeed, we identified dysregulated chemokine receptor expression on ILC2 in COVID-19 patients, specifically CXCR3 which was reduced in the patients as compared with controls (Fig. 3E). PCA analysis provided an illustration of these findings by segregating the ILC2 data from patients and controls based on CD69 expression for patients and chemokine receptors as well as CD62L and CD45RA for the controls (Fig. 3F). The latter two were, however, not statistically different between the groups (Fig. 3E).

ILCp showed an activated profile in COVID-19 patients, with increased CD69 and HLA-DR expression as compared with controls. Additionally, CXCR3 was decreased on ILCp in COVID-19 patients as compared with controls (Fig. 3G). Although PCA analysis did not separate the ILCp phenotype from patients and controls clearly, there was a tendency towards separation of ILCp from controls on the basis of CD62L and CD45RA (Fig. 3H). These two markers showed reduced tendency but were not statistically significant in the manual gating (Fig. 3G).

Overall, these findings suggest that the ILCs remaining in the circulation of COVID-19 patients are activated and show an altered expression of chemokine receptors, particularly a decrease in CXCR3, possibly reflecting alterations in CXCR3-ligand dependent ILC recruitment to tissues.

Uniform Manifold Approximation and Projection (UMAP) analysis confirmed and extended the findings obtained by manual gating in an unsupervised manner. As expected from manual gating, clear differences in ILC composition between controls and COVID-19 patients were observed when overlaying each group to the UMAP containing all concatenated ILC events (Fig. 4A, S2B). Phenograph clustering yielded a total of 14 distinct clusters across patients and controls (Fig. 4B). Based on the relative expression of CRTH2, CD117 and CXCR3, clusters were identified as ILC1, ILC2 or ILCp (Fig. 4C-D). Thereafter, manually defined ILC subset gates that were overlayed into the UMAP (Fig. 4E) showed a similar spatial distribution as the Phenograph-defined clusters belonging to each ILC subset, confirming the precision of the manual gating relative to the unsupervised analysis (Fig. 4B).

**Figure 4.**
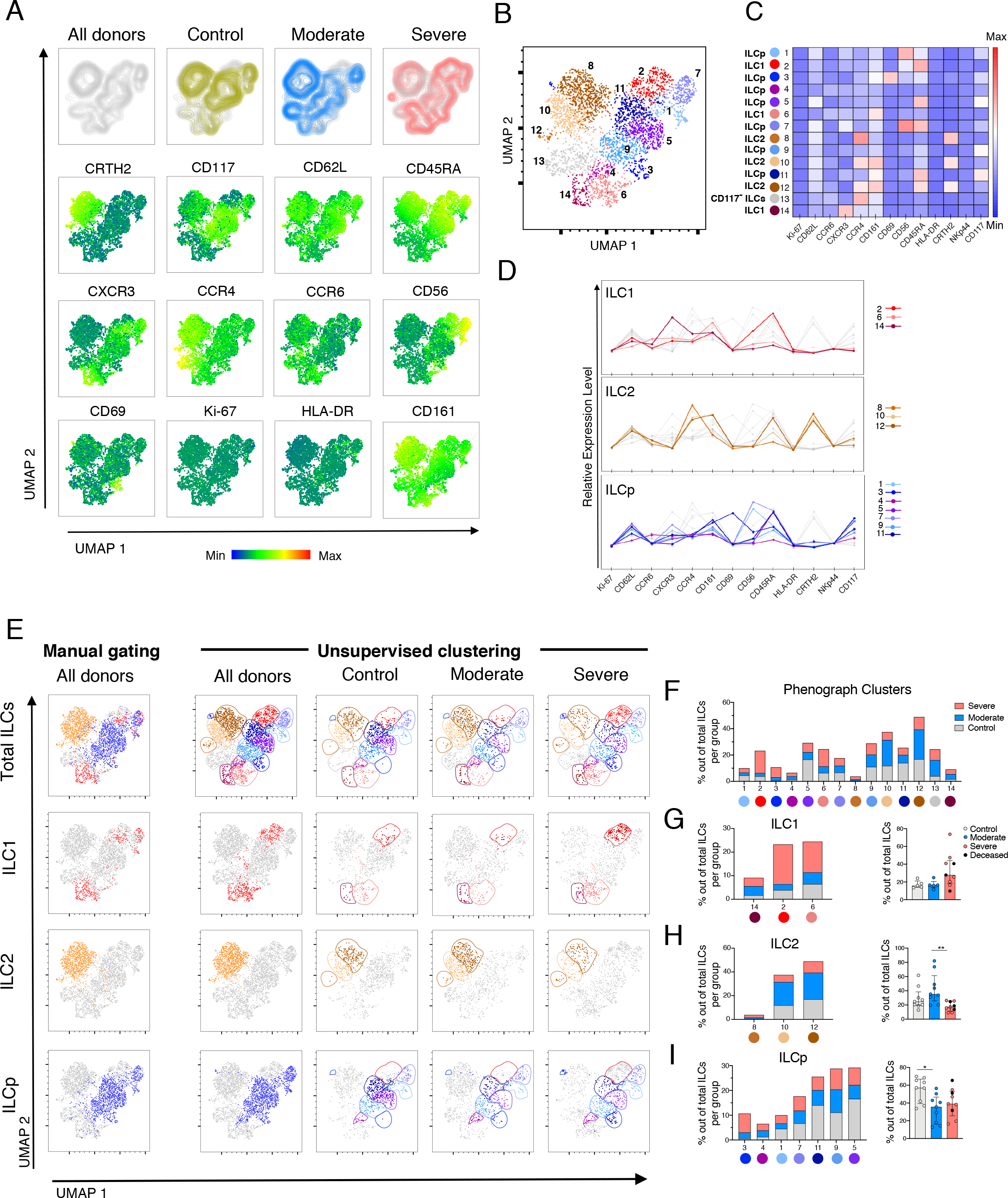
Dimensionality reduction analysis of ILCs in the peripheral blood of COVID-19 patients distinguishes moderate and severe patients. **(A)** Top row: UMAP of total ILCs from controls and COVID-19 patients (All donors) and overlaid by patient groups: of control donors (yellow), moderate COVID-19 patients (blue) and severe COVID-19 patients (red) (from left to right). Middle and bottom rows: UMAP (All donors) colored according to the fluorescence intensity expression (median) of the indicated phenotypic markers. **(B)** UMAP of the total ILCs overlaid with the 14 clusters identified by Phenograph. **(C)** Heatmap displaying the median of expression of the indicated markers for each Phenograph clusters. Each cluster was assigned an ILC subset identity (ILC1, ILC2, ILCp and CD117^-^ ILC) based on the heatmap and the relative expression levels in (D). **(D)** Relative expression level of markers in the Phenograph clusters grouped by ILC subsets (ILC1, ILC2 and ILCp). Grey lines in each graph are the rest of clusters not belonging to the ILC subset depicted. **(E)** Far left column: manually defined gates of total ILCs and ILC subsets ILC1, ILC2 and ILCp overlayed on the All donors UMAP in (A). Right columns: UMAP of total ILCs overlaid with the 14 ILCs clusters identified by Phenograph and displayed according to patient group (columns) and ILCs subsets (rows). Colors used for the clusters correspond to the colors used in Fig. 4B-D. **(F)** Stacked bar graph of the percentage of all Phenograph-identified clusters out of total ILCs in controls (grey), moderate (blue) and severe COVID-19 patients (red). **(G)** Left: stacked bar graph of the percentage of the Phenograph-identified clusters belonging to ILC1 out of total ILCs in controls (grey), moderate (blue) and severe (red) COVID-19 patients. Right: Percentage of the sum of events corresponding to the ILC1 Phenograph-identified clusters (14,2,6) out of total ILCs, in controls (n=5), moderate (n=5), and severe COVID-19 patients (n=9). **(H)** Left: stacked bar graph of the percentage of the Phenograph-identified clusters belonging to ILC2 out of total ILCs in controls, moderate, and severe COVID-19 patients. Right: Percentage of the sum of events corresponding to the ILC2 Phenograph-identified clusters (8,10,12) out of total ILCs, in controls (n=9), moderate (n=9), and severe COVID-19 patients (n=9). **(I)** Left: stacked bar graph of the percentage of the Phenograph-identified clusters belonging to ILCp out of total ILCs in controls, moderate, and severe COVID-19 patients. Right: percentage of the sum of events corresponding to the ILCp Phenograph-identified clusters (3,4,1,7,11,9,5) out of total ILCs, in controls (n=9), moderate (n=9), and severe COVID-19 patients (n=9). **(G-I)** Patients with less than 10 events per ILC subsets (defined by Phenograph-identified clusters) were excluded from analysis. Statistical differences were tested using the Kruskal-Wallis test followed by Dunn’s multiple comparisons test. Bar graphs are shown as median ± IQR, *p < 0.05, ** p < 0.01.

Next, we assessed each Phenograph-defined cluster. Validating the manual gating, CCR4^+^ ILC1 (cluster 14) was uniquely present in COVID-19 patients, while two additional CCR4^-/lo^ ILC1 clusters (2 and 6) were overrepresented in severe COVID-19 patients (Fig. 4F, G). For ILC2, cluster 8, containing CCR4^hi^ ILC2, was only present in COVID-19 patients and other two CCR6^+^ ILC2 clusters (10, 12) were accumulated in moderate patients (Fig. 4F, H). Corroborating again the findings from manual gating (Fig. 3), cluster 3, uniquely present in COVID-19 patients, corresponded to activated (CD69^+^) ILCp lacking CXCR3 (Fig. 4F, I). Furthermore, three ILCp clusters (1, 5, and 7) with an immature phenotype (CD45RA^+/hi^/CD62L^+/hi^) were reduced in COVID-19 patients as compared with controls (Fig. 4F, I), in support of the data obtained by manual gating (Fig. 3).

Altogether, the unsupervised analysis revealed clusters specifically accumulated in COVID-19 patients compared to controls and the presence of specific ILC phenotypes that associated to the disease severity. Importantly, several of the findings agreed with those obtained by manual gating. Specifically, we confirmed an increase in the relative frequency of CD69^+^ ILCp and a decrease in CXCR3^+^ ILCp in both moderate and severe COVID-19, an increase in ILC2 percentage specifically in moderate COVID-19 and an increased CCR4 expression among ILC1 in the patients.

To analyze for potential factors involved in the activation and potential recruitment of ILCs to tissues in the COVID-19 patients, we examined correlations between ILCs and a wide array of soluble serum factors (Fig. S5A-B). Based on our findings on altered frequencies of CXCR3^+^ and CD69^+^ ILCs in COVID-19 (Fig. 3A), we focused on selected chemokines (CCL20, CXCL10 and CXCL11) and factors previously reported to be increased in COVID-19 or other viral respiratory disease: IL-6, IL-10, IL18R1 and PD-L1^7–9,37–42^. Interestingly, we found that the percentage of activated (CD69^+^) total ILCs and activated ILCp positively correlated with serum IL-6 levels in the COVID-19 patients (Fig. 5A). Additionally, the relative levels of the cytokine IL-10 also positively correlated with the levels of CD69^+^ ILCs in the COVID-19 patients (Fig. 5B). Moreover, the percentage of CD69^+^ total ILCs and ILCp positively correlated with CXCL10 levels in the COVID-19 patients (Fig. 5C), suggesting that this chemokine is related to the increase in the percentage of activated ILCs that remain in the circulation of COVID-19 patients.

**Figure 5.**
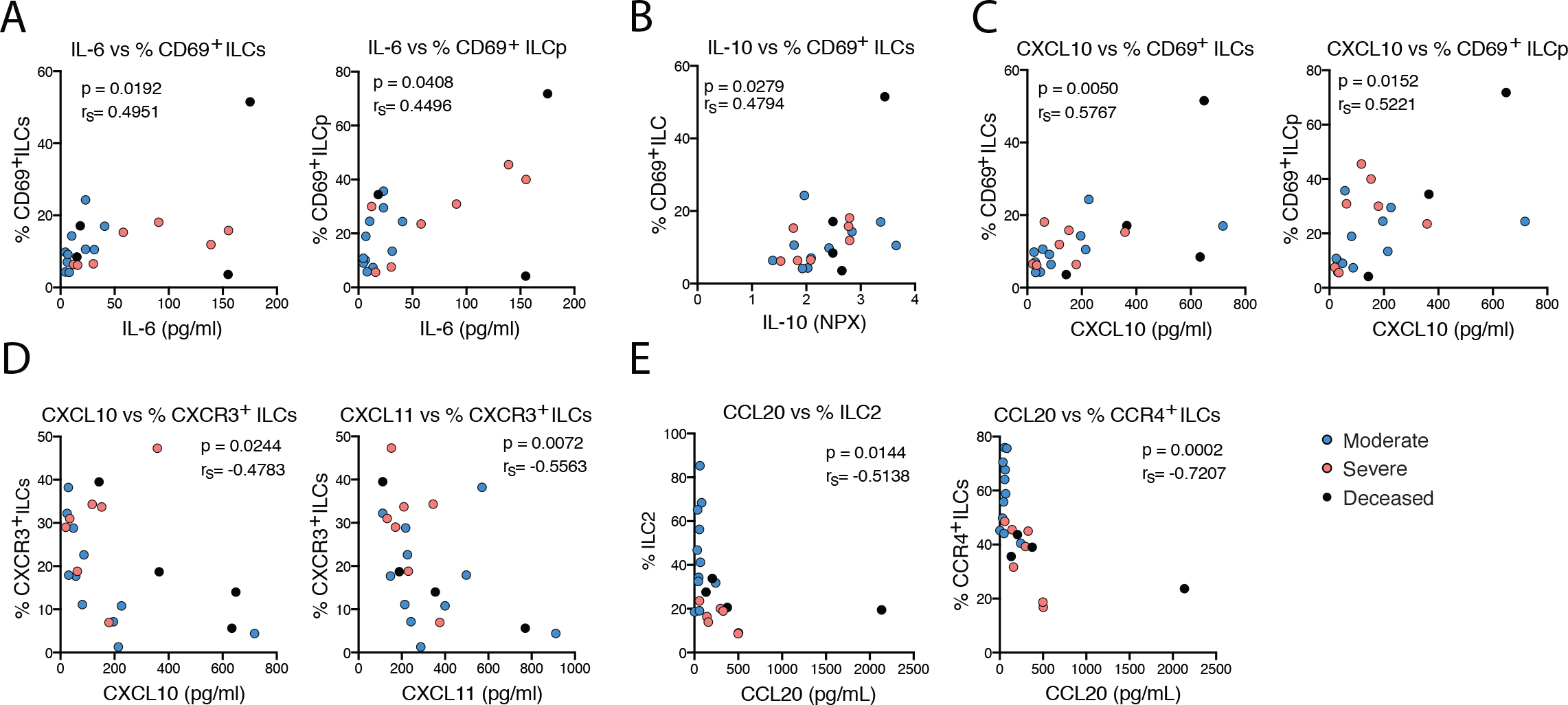
Activation status and homing profile of peripheral blood ILCs associate with inflammation markers in COVID-19 patients. Spearman correlations between **(A)** serum IL-6 levels (pg/ml) and the percentage of CD69^+^ ILC and CD69^+^ ILCp; **(B)** serum IL-10 relative levels and the percentage of CD69^+^ ILC in COVID-19 patients; **(C)** serum CXCL10 levels (pg/ml) and the percentage of CD69^+^ ILC and CD69^+^ ILCp; **(D)** serum CXCL10 and CXCL11 levels (pg/ml) and the percentage of CXCR3^+^ ILCs; **(E)** serum CCL20 levels (pg/ml) and the percentage of ILC2 and CCR4^+^ ILCs. IL-6, CXCL10, CXCL11 and CCL20 serum absolute levels were measured with Magnetic Luminex Screening assay, and IL-10 relative levels with a proximity extension assay, where data is shown as normalized protein expression (NPX). Blue circles: moderate COVID-19 patients (n=11); red circles: severe COVID-19 patients (n=11); black circles: deceased severe COVID-19 patients (n=4). p < 0.05 was considered statistically significant. r_s:_ Spearman’s rank correlation coefficient.

Furthermore, indicative of dysregulated ILC tissue migration in COVID-19, we observed a negative correlation of CXCL10 and CXCL11 levels (CXCR3 ligands), with the percentage of CXCR3^+^ ILCs in COVID-19 patients (Fig. 5D). Additionally, the percentage of CCR4^+^ total ILCs and ILC2 negatively correlated with CCL20 (ligand for CCR6) levels. The latter correlation is in line with the high expression of CCR6 on circulating ILC2 (ref. 17) (Fig. 5E).

Of further interest, IL18R1 and PD-L1, involved in type 1-inflammation^40,43^ negatively correlated with the level of CD45RA^+^ ILCp, suggesting that the inflammatory status of COVID-19 patients may promote maturation, depletion and/or tissue migration of ILCp in the systemic circulation (Fig. S5C).

We next sought to integrate our flow cytometric data with the clinical and laboratory findings from the same patients. Clinical information and laboratory measurements (summarized in Tables 1-2) integrated in a PCA analysis clustered the patients on the basis of disease severity (Fig. 6A) with several hematological and biochemical factors particularly driving this separation (Fig. 6B).

**Figure 6.**
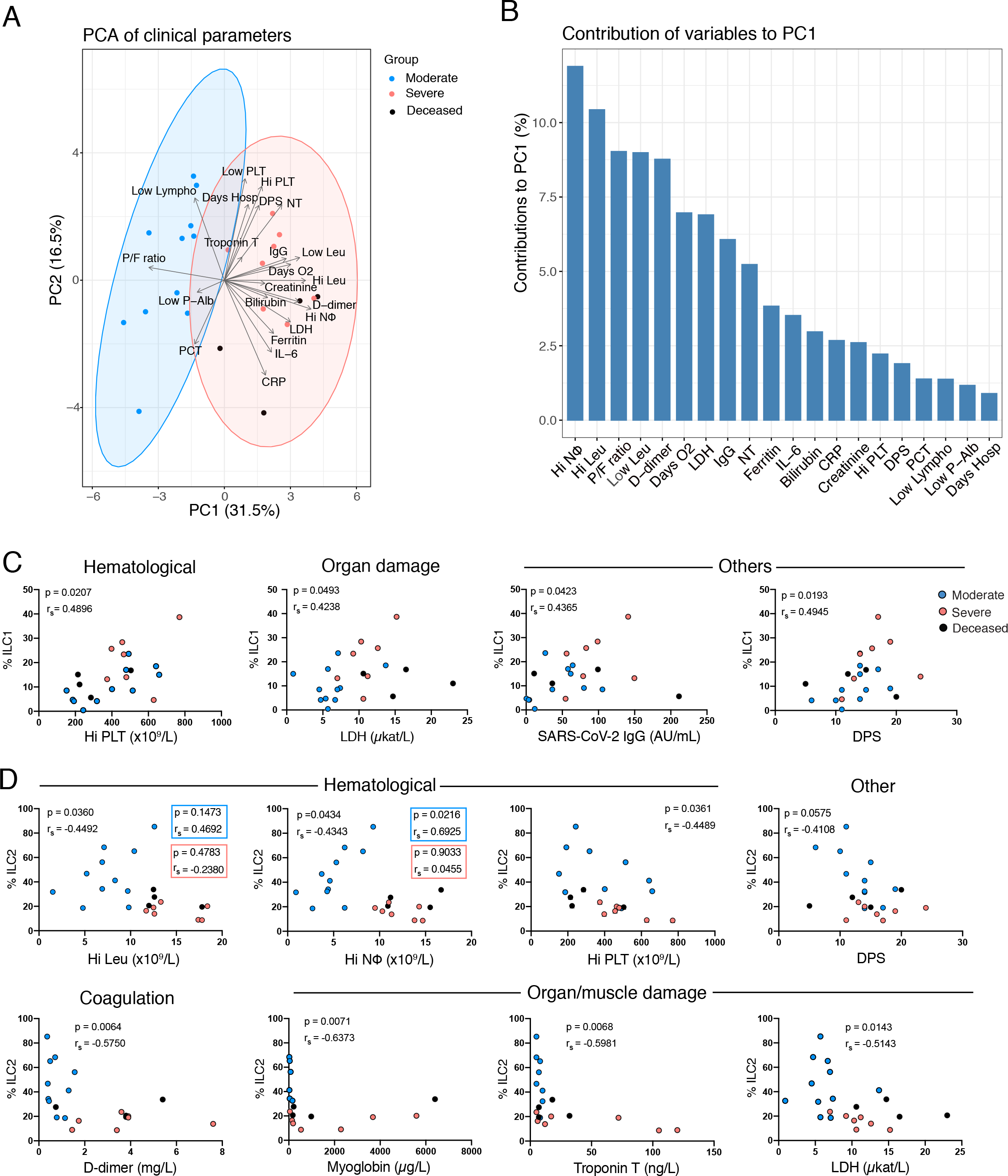
Peripheral blood ILC subsets associate with biochemical and hematological parameters that reflect COVID-19 severity. **(A)** Principal Component Analysis (PCA) of COVID-19 patients displaying the distribution and segregation of COVID-19 patients according to clinical and laboratory parameters. **(B)** Bar plot showing the percentage contribution of each clinical or laboratory parameter to principal component 1 (PC1). **(C)** Correlation plots between percentage of ILC1 and the indicated hematological (Hi PLT), organ damage (LDH) and other parameters (SARS-CoV-2 IgG and DPS) in COVID-19 patients (moderate n=11; severe n=11). **(D)** Correlation plots between percentage of ILC2 and the indicated hematological (Hi Leu, Hi NΦ, Hi PLT and DPS), coagulation (D-dimer), organ damage (myoglobin, troponin and LDH) and other (SARS-CoV-2 IgG and DPS) parameters in COVID-19 patients (moderate n=11;severe n=11). Hi NΦ: highest neutrophil count +/-24h from sampling; PF ratio: PaO2/FiO2 ratio (mm Hg) at sampling; Hi PLT: highest platelet count before sampling (b.s); IL-6: IL-6 levels at the time of sampling; NT: neutralizing antibody titers at sampling; CRP: highest C-reactive protein +/-24h; LDH: highest lactate dehydrogenase b.s; PCT: highest procalcitonin +/-24h; Days O_2_: days of oxygen treatment; Low Lympho: Lowest lymphocyte count +/-24h; Low P-Alb: lowest P-Albumin +/-24h; Hi Leu: highest leukocyte count +/-24h; Low Leu: Lowest leukocyte count +/-24h; D-dimer: highest D-dimer level +/-24h; Ferritin: highest ferritin level +/-24h; SARS-CoV-2 IgG: SARS-CoV-2 IgG antibodies (AU/mL); DPS: days post symptom debut to sampling; Days Hosp: days of hospitalization until sampling. Spearman’s rank correlation test was applied for assessing correlations between variables.

We searched for potentially relevant correlations between ILCs and the clinical and laboratory parameters. The frequencies of ILC1 and ILC2 correlated with several of the measured parameters, whereas no correlations were found for ILCp frequencies (Fig. S3B). The percentage of ILC1 positively correlated with LDH levels, which was significantly increased in severe COVID-19 patients (Fig. 6C, Fig. S1). ILC1 frequencies also positively correlated with parameters that did not differ between the two COVID-19 patient groups, i.e. platelet counts, serum SARS-CoV-2 IgG levels and days post symptom debut (Fig. 6C). The percentage of ILC2, on the other hand, negatively correlated with the leukocyte, neutrophil and platelet count (Fig. 6D). Interestingly, high neutrophil levels, which have been described as partial predictors of disease severity^44^, also contributed heavily to the separation between patient groups in the current study (Fig. 6A-B). Of note, neutrophils have been found to inhibit ILC2 function, thus preventing allergic airway inflammation^45^. Additionally, the relative frequency of ILC2 correlated negatively with levels of D-dimer, a coagulation factor that has been suggested as a systemic biomarker of disease severity in COVID-19 (Fig. 6D)^44,46^. Finally, we observed a negative correlation between ILC2 frequencies and organ/muscle damage markers (myoglobin, troponin T, LDH) and the number of days since symptom debut (Fig. 6D). These findings suggest that among COVID-19 patients, elevated frequencies of ILC1 and decreased frequencies of ILC2 are hallmarks of COVID-19 patients with a clinical profile associated with severe disease.

## DISCUSSION

We report unphysiologically-reduced levels of ILCs in the circulation of COVID-19 patients, both in percentage and absolute counts, in line with a recent report^47^, and the overall reduction of peripheral blood lymphocytes described in COVID-19^6,38,48,49^. In addition to the overall ILC depletion, we found major changes in the residual circulating ILC compartment in COVID-19 patients, including altered frequencies of ILC subsets, increased activation and dysregulated migration receptor expression. Our data imply that the circulating ILCs are activated and have differentiated and/or are differentially recruited to tissues where they may contribute to the anti-viral defense.

Noteworthy, we found increased circulating ILC2 frequencies in moderate but not severe COVID-19 patients, suggesting that ILC2 may be differentially regulated dependent on severity of disease. Supporting this, the relative frequency of ILC2 in COVID-19 patients correlated negatively with the coagulation factor D-dimer, previously shown to be associated with the development of severe disease^44,46^ as well as organ/muscle damage markers, suggesting that low ILC2 levels in COVID-19 patients could be indicative of a more severe disease outcome. Indeed, ILC2 have been shown to have a prominent role in lung tissue repair during influenza A infection in mice^22^. This was achieved through production of the epidermal growth factor related protein amphiregulin, also shown to be involved in promotion of regulatory T cells and cell survival in hepatitis C virus infection^50,51^. The role for ILC2-derived amphiregulin in lung tissue repair in humans is unknown and deserves further exploration in COVID-19.

In contrast to ILC2, ILCp frequencies were diminished in COVID-19 patients as compared with controls, suggesting their migration to the site of infection or differentiation into mature ILC subsets in the circulation. This has previously been described in an adoptive transfer mouse model^18^. In support of this hypothesis, both ILC2 and ILCp showed an activated phenotype, based on increased frequencies of CD69^+^ and/or HLA-DR^+^ cells. This is in line with the recent discovery of increased frequencies of CD69^+^ NK cells in COVID-19^52^. Interestingly, in addition to its known role as an early activation marker, CD69 is also a marker of tissue residency^53^. Thus, the presence of the CD69-expressing ILCs in circulation could also indicate retrograde migration from tissue to circulation after local tissue activation, as recently shown for resident memory T cells^54^. Furthermore, we observed that the CD69-expressing cells in the circulation of COVID-19 patients positively correlated with the levels of IL-6 and IL-10, suggesting that the inflammatory status in these patients could be the driver of ILC activation and/or recirculation. Additionally, these CD69-expressing cells positively correlated with the chemokine CXCL10 in patients. In contrast, CXCR3^+^ ILCs negatively correlated with CXCR3 ligands CXCL10 and CXCL11, potentially indicating a CXCR3-dependent tissue migration of ILCs in COVID-19. CXCR3^+^ ILCs includes IFN-γ producing ILC1 (ref. 11,20), which are accumulated in acute and chronic intestinal inflammation in humans^11^. While ILC1 are essential for clearance of cytomegalovirus in mice^55^, the role for ILC1 in human anti-viral immunity, including COVID-19, remains obscure and requires further exploration.

Lastly, the phenotypic characterization of circulating ILCs performed in this study would benefit from additional analyses delineating the functionality of ILCs as well as the impact of the tissue microenvironment to their phenotype and function. Of special interest would be the study of ILCs from tissue resident sites, such as lung and intestine, which have been described as main sites of SARS-CoV-2 infection and shedding^56–59^. We have previously reported increased ILC2 frequencies in the intestine of patients with inflammatory bowel disease^17^ and in the lungs of patients with asthma^60^, likely due to their influx from the circulation. In line with this, it is possible that the lower ILC frequencies observed in severe as compared with moderate COVID-19 patients could indicate potential trafficking of ILC2 to inflamed infection sites. To address whether elevated levels of circulating ILC2 constitute a correlate of protection in COVID-19 patients, further studies at the tissue level are required.

In summary, this study provides a phenotypic characterization of circulating ILCs of COVID-19 patients. We identified that altered frequencies of ILCs correlate with clinical and laboratory parameters linked to COVID-19 disease severity. Lastly, to the best of our knowledge, this is the first comprehensive study of ILCs in the context of a human respiratory viral infection. Thus, these data pave the way for a better understanding of the role of this innate immune cell compartment in other viral infectious diseases.

## Data Availability

Curated flow cytometry data is available for exploration via the Karolinska COVID-19 Immune
Atlas (homepage currently under construction). Other data are available upon request to the corresponding author.

## AUTHOR CONTRIBUTION

M.G.: conceptualization, data curation, formal analysis, investigation, methodology, and writing-original draft; E.K.: conceptualization, data curation, formal analysis, investigation, methodology, and writing-original draft; A.C.G.: conceptualization, investigation, methodology, and writing-review and editing; T.P.: conceptualization, methodology, and writing-review and editing; L.M.P.M.: Conceptualization, data curation, formal analysis, and writing-review and editing; K.T.M.: conceptualization, methodology, and writing-review and editing; W.C.: conceptualization, methodology, and writing-review and editing; R.V.: conceptualization, resources, and writing-review and editing; I.F.: conceptualization, resources, and writing-review and editing; H.G.L.: conceptualization, funding acquisition, and writing-review and editing; N.K.B: conceptualization and writing-review and editing; E.F.: methodology and writing-review and editing; O.R.: conceptualization, methodology, and writing-review and editing; L.I.E.: conceptualization and writing-review and editing; A.S.: conceptualization and writing-review and editing; S.A.: conceptualization, methodology, and writing-review and editing; K.S.: conceptualization, methodology, investigation, and writing-review and editing; S.G.R.: conceptualization, investigation, and writing-review and editing; J.K.: conceptualization, investigation, supervision, funding acquisition, and writing-original draft; J.M.: conceptualization, investigation, supervision, funding acquisition, and writing-original draft; Karolinska KI/K COVID-19 Study Group: conceptualization, methodology, and writing-review and editing.

## ACKNOWLEDGEMENTS

We express our sincere gratitude to all the patients and their families as well as the clinical personnel that helped to carry out the study as part of the Karolinska KI/K COVID-19 Immune Atlas.

## REFERENCES

1. Alijotas-Reig J, Esteve-Valverde E, Belizna C, et al. Immunomodulatory therapy for the management of severe COVID-19. Beyond the anti-viral therapy: A comprehensive review. Autoimmun Rev 2020; 19(7). doi:10.1016/j.autrev.2020.102569

2. Riva L, Yuan S, Yin X, et al. Discovery of SARS-CoV-2 antiviral drugs through large-scale compound repurposing. Nature 2020; 586(7827): 113–119. doi:10.1038/s41586-020-2577-1

3. Thanh Le T, Andreadakis Z, Kumar A, et al. The COVID-19 vaccine development landscape. Nat Rev Drug Discov 2020; 19(5): 305–306. doi:10.1038/d41573-020-00073-5

4. Zhou P, Yang X Lou, Wang XG, et al. A pneumonia outbreak associated with a new coronavirus of probable bat origin. Nature 2020; 579(7798): 270–273. doi:10.1038/s41586-020-2012-7

5. Chen G, Wu D, Guo W, et al. Clinical and immunological features of severe and moderate coronavirus disease 2019. J Clin Invest 2020; 130(5): 2620–2629. doi:10.1172/JCI137244

6. Huang C, Wang Y, Li X, et al. Clinical features of patients infected with 2019 novel coronavirus in Wuhan, China. Lancet 2020; 395(10223): 497–506. doi:10.1016/S0140-6736(20)30183-5

7. Azkur AK, Akdis M, Azkur D, et al. Immune response to SARS-CoV-2 and mechanisms of immunopathological changes in COVID-19. Allergy Eur J Allergy Clin Immunol 2020; 75(7): 1564–1581. doi:10.1111/all.14364

8. Mahmudpour M, Roozbeh J, Keshavarz M, Farrokhi S, Nabipour I. COVID-19 cytokine storm: The anger of inflammation. Cytokine 2020; 133: 155151. doi:10.1016/j.cyto.2020.155151

9. Mehta P, McAuley DF, Brown M, Sanchez E, Tattersall RS, Manson JJ. COVID- 19: consider cytokine storm syndromes and immunosuppression. Lancet 2020; 395(10229): 1033–1034. doi:10.1016/S0140-6736(20)30628-0

10. Vivier E, Artis D, Colonna M, et al. Innate Lymphoid Cells: 10 Years On. Cell. 2018; 174(5): 1054–1066. doi:10.1016/j.cell.2018.07.017

11. Bernink JH, Peters CP, Munneke M, et al. Human type 1 innate lymphoid cells accumulate in inflamed mucosal tissues. Nat Immunol 2013; 14(3): 221–229. doi:10.1038/ni.2534

12. Klose CSN, Kiss EA, Schwierzeck V, et al. A T-bet gradient controls the fate and function of CCR6-RORγt + innate lymphoid cells. Nature 2013; 494(7436): 261–265. doi:10.1038/nature11813

13. Moro K, Yamada T, Tanabe M, et al. Innate production of TH 2 cytokines by adipose tissue-associated c-Kit+ Sca-1+ lymphoid cells. Nature 2010; 463(7280): 540–544. doi:10.1038/nature08636

14. Neill DR, Wong SH, Bellosi A, et al. Nuocytes represent a new innate effector leukocyte that mediates type-2 immunity. Nature 2010; 464(7293): 1367–1370. doi:10.1038/nature08900

15. Price AE, Liang HE, Sullivan BM, et al. Systemically dispersed innate IL-13- expressing cells in type 2 immunity. Proc Natl Acad Sci U S A 2010; 107(25): 11489–11494. doi:10.1073/pnas.1003988107

16. Satoh-Takayama N, Vosshenrich CAJ, Lesjean-Pottier S, et al. Microbial Flora Drives Interleukin 22 Production in Intestinal NKp46+ Cells that Provide Innate Mucosal Immune Defense. Immunity 2008; 29(6): 958–970. doi:10.1016/j.immuni.2008.11.001

17. Forkel M, VanTol S, Höög C, Michaëlsson J, Almer S, Mjösberg J. Distinct alterations in the composition of mucosal innate lymphoid cells in newly diagnosed and established Crohn’s disease and ulcerative colitis. J Crohn’s Colitis 2019; 13(1): 67–78. doi:10.1093/ecco-jcc/jjy119

18. Lim AI, Li Y, Lopez-Lastra S, et al. Systemic Human ILC Precursors Provide a Substrate for Tissue ILC Differentiation. Cell 2017; 168(6): 1086-1100.e10. doi:10.1016/j.cell.2017.02.021

19. Mjösberg J, Bernink J, Golebski K, et al. The Transcription Factor GATA3 Is Essential for the Function of Human Type 2 Innate Lymphoid Cells. Immunity 2012; 37(4): 649–659. doi:10.1016/j.immuni.2012.08.015

20. Roan F, Stoklasek TA, Whalen E, et al. CD4 + Group 1 Innate Lymphoid Cells (ILC) Form a Functionally Distinct ILC Subset That Is Increased in Systemic Sclerosis. J Immunol 2016; 196(5): 2051–2062. doi:10.4049/jimmunol.1501491

21. Simoni Y, Fehlings M, Kløverpris HN, et al. Human Innate Lymphoid Cell Subsets Possess Tissue-Type Based Heterogeneity in Phenotype and Frequency. Immunity 2017; 46(1): 148–161. doi:10.1016/j.immuni.2016.11.005

22. Monticelli LA, Sonnenberg GF, Abt MC, et al. Innate lymphoid cells promote lung-tissue homeostasis after infection with influenza virus. Nat Immunol 2011; 12(11): 1045–1054. doi:10.1038/ni.2131

23. Li BWS, de Bruijn MJW, Lukkes M, et al. T cells and ILC2s are major effector cells in influenza-induced exacerbation of allergic airway inflammation in mice. Eur J Immunol 2019; 49(1): 144–156. doi:10.1002/eji.201747421

24. Kløverpris HN, Kazer SW, Mjösberg J, et al. Innate Lymphoid Cells Are Depleted Irreversibly during Acute HIV-Infection in the Absence of Viral Suppression. Immunity 2016; 44(2): 391–405. doi:10.1016/j.immuni.2016.01.006

25. Beigel JH, Tomashek KM, Dodd LE, et al. Remdesivir for the Treatment of Covid-19 — Preliminary Report. N Engl J Med 2020; 383(10): 992–993. doi:10.1056/nejmoa2007764

26. Coronavirus disease (COVID-19). https://www.who.int/emergencies/diseases/novel-coronavirus-2019.

27. Wei R, Wang J, Jia E, Chen T, Ni Y, Jia W. GSimp: A Gibbs sampler based left- censored missing value imputation approach for metabolomics studies. PLoS Comput Biol 2018; 14(1). doi:10.1371/journal.pcbi.1005973

28. R Core Team. R: A language and environment for statistical computing. R Foundation for Statistical Computing, Vienna, Austria. https://www.r-project.org/.

29. Sekine T, Perez-Potti A, Rivera-Ballesteros O, et al. Robust T cell immunity in convalescent individuals with asymptomatic or mild COVID-19. Cell 2020; 183(1): 158-168.e14. doi:10.1101/2020.06.29.174888

30. Varnaitė R, García M, Glans H, et al. Expansion of SARS-CoV-2- Specific Antibody-Secreting Cells and Generation of Neutralizing Antibodies in Hospitalized COVID-19 Patients. J Immunol. 2020. Epub ahead of print. doi:10.4049/jimmunol.2000717

31. Kassambara, A. and Mundt F. factoextra: Extract and Visualize the Results of Multivariate Data Analyses. R package version 1.0.7. https://cran.r-project.org/web/packages/factoextra/index.html.

32. Lê S, Josse J, Husson F. FactoMineR: An R package for multivariate analysis. J Stat Softw 2008; 25(1): 1–18. doi:10.18637/jss.v025.i01

33. Neuwirth E. RColorBrewer: ColorBrewer Palettes. R package version 1.1-2https://cran.r-project.org/web/packages/RColorBrewer/index.html.

34. Wickham H. ggplot2: Elegant Graphics for Data Analysis. https://ggplot2.tidyverse.org/.

35. Cheronet, O., and Finarelli JA. WaverR: Data Estimation using Weighted Averages of Multiple Regressions. https://cran.r-project.org/web/packages/WaverR/index.html.

36. Yudanin NA, Schmitz F, Flamar AL, et al. Spatial and Temporal Mapping of Human Innate Lymphoid Cells Reveals Elements of Tissue Specificity. Immunity 2019; 50(2): 505-519.e4. doi:10.1016/j.immuni.2019.01.012

37. Liu F, Li L, Xu M Da, et al. Prognostic value of interleukin-6, C-reactive protein, and procalcitonin in patients with COVID-19. J Clin Virol 2020; 127: 104370. doi:10.1016/j.jcv.2020.104370

38. Zhang X, Tan Y, Ling Y, et al. Viral and host factors related to the clinic outcome of the SARS-CoV-2 infection. Nature 2020; 583(7816): 437–440. doi:10.1038/s41586-020-2355-0

39. Maleki KT, García M, Iglesias A, et al. Serum Markers Associated with Severity and Outcome of Hantavirus Pulmonary Syndrome. J Infect Dis 2019; 219(11): 1832–1840. doi:10.1093/infdis/jiz005

40. Schönrich G, Raftery MJ. The PD-1/PD-L1 axis and virus infections: A delicate balance. Front Cell Infect Microbiol 2019; 9: 207. doi:10.3389/fcimb.2019.00207

41. Zalinger ZB, Elliott R, Weiss SR. Role of the inflammasome-related cytokines Il- 1 and Il-18 during infection with murine coronavirus. J Neurovirol 2017; 23(6): 845–854. doi:10.1007/s13365-017-0574-4

42. Harker JA, Godlee A, Wahlsten JL, et al. Interleukin 18 Coexpression during Respiratory Syncytial Virus Infection Results in Enhanced Disease Mediated by Natural Killer Cells. J Virol 2010; 84(8): 4073–4082. doi:10.1128/jvi.02014-09

43. Dinarello CA, Novick D, Kim S, Kaplanski G. Interleukin-18 and IL-18 binding protein. Front Immunol 2013; 4: 289. doi:10.3389/fimmu.2013.00289

44. Velavan TP, Meyer CG. Mild versus severe COVID-19: Laboratory markers. International Journal of Infectious Diseases 2020; 95: 304–307. doi:10.1016/j.ijid.2020.04.061

45. Patel DF, Peiró T, Bruno N, et al. Neutrophils restrain allergic airway inflammation by limiting ILC2 function and monocyte-dendritic cell antigen presentation. Sci Immunol 2019;4(41): 7006. doi:10.1126/sciimmunol.aax7006

46. Zhou F, Yu T, Du R, et al. Clinical course and risk factors for mortality of adult inpatients with COVID-19 in Wuhan, China: a retrospective cohort study. Lancet 2020; 395(10229): 1054–1062. doi:10.1016/S0140-6736(20)30566-3

47. Kuri-Cervantes L, Pampena MB, Meng W, et al. Immunologic perturbations in severe COVID-19/SARS-CoV-2 infection. bioRxiv [Preprint]. 2020; doi:10.1101/2020.05.18.101717

48. Qin C, Zhou L, Hu Z, et al. Dysregulation of Immune Response in Patients With Coronavirus 2019 (COVID-19) in Wuhan, China. Clin Infect Dis 2020; 71(15): 762–768. doi:10.1093/cid/ciaa248

49. Wang F, Nie J, Wang H, et al. Characteristics of Peripheral Lymphocyte Subset Alteration in COVID-19 Pneumonia. J Infect Dis 2020; 221(11): 1762–1769. doi:10.1093/infdis/jiaa150

50. Pei R, Chen H, Lu L, et al. Hepatitis C virus infection induces the expression of amphiregulin, a factor related to the activation of cellular survival pathways and required for efficient viral assembly. J Gen Virol 2011; 92(10): 2237–2248. doi:10.1099/vir.0.032581-0

51. Yuan CH, Sun XM, Zhu CL, et al. Amphiregulin activates regulatory T lymphocytes and suppresses CD8+ T cell-mediated anti-tumor response in hepatocellular carcinoma cells. Oncotarget 2015; 6(31): 32138–32153. doi:10.18632/oncotarget.5171

52. Maucourant C, Filipovic I, Ponzetta A, et al. Natural killer cell immunotypes related to COVID-19 disease severity. Sci Immunol 2020; 5(50): eabd6832. doi:10.1126/sciimmunol.abd6832

53. Kumar B V, Ma W, Miron M, et al. Human Tissue-Resident Memory T Cells Are Defined by Core Transcriptional and Functional Signatures in Lymphoid and Mucosal Sites. Cell Reports 2017; 20(12): 2921–2934. doi:10.1016/j.celrep.2017.08.078

54. Fonseca R, Beura LK, Quarnstrom CF, et al. Developmental plasticity allows outside-in immune responses by resident memory T cells. Nat Immunol 2020; 21(4): 412–421. doi:10.1038/s41590-020-0607-7

55. Weizman O El, Adams NM, Schuster IS, et al. ILC1 Confer Early Host Protection at Initial Sites of Viral Infection. Cell 2017; 171(4): 795-808.e12. doi:10.1016/j.cell.2017.09.052

56. Cheung KS, Hung IFN, Chan PPY, et al. Gastrointestinal Manifestations of SARS-CoV-2 Infection and Virus Load in Fecal Samples From a Hong Kong Cohort: Systematic Review and Meta-analysis. Gastroenterology 2020; 159(1): 81–95. doi:10.1053/j.gastro.2020.03.065

57. Lamers MM, Beumer J, van der Vaart J, et al. SARS-CoV-2 productively infects human gut enterocytes. Science 2020; 369(6499): 50–54. doi:10.1126/science.abc1669

58. Schaefer IM, Padera RF, Solomon IH, et al. In situ detection of SARS-CoV-2 in lungs and airways of patients with COVID-19. Mod Pathol 2020. Epub ahead of print. doi:10.1038/s41379-020-0595-z

59. Xiao F, Tang M, Zheng X, Liu Y, Li X, Shan H. Evidence for Gastrointestinal Infection of SARS-CoV-2. Gastroenterology 2020; 158(6): 1831-1833.e3. doi:10.1053/j.gastro.2020.02.055

60. Winkler C, Hochdörfer T, Israelsson E, et al. Activation of group 2 innate lymphoid cells after allergen challenge in asthmatic patients. J Allergy Clin Immunol 2019; 144(1): 61-69.e7. doi:10.1016/j.jaci.2019.01.027

