## Supplementary Material for "Innate lymphoid cell composition associates with COVID-19 disease severity"

**García *et al*. Supplementary figures and tables**

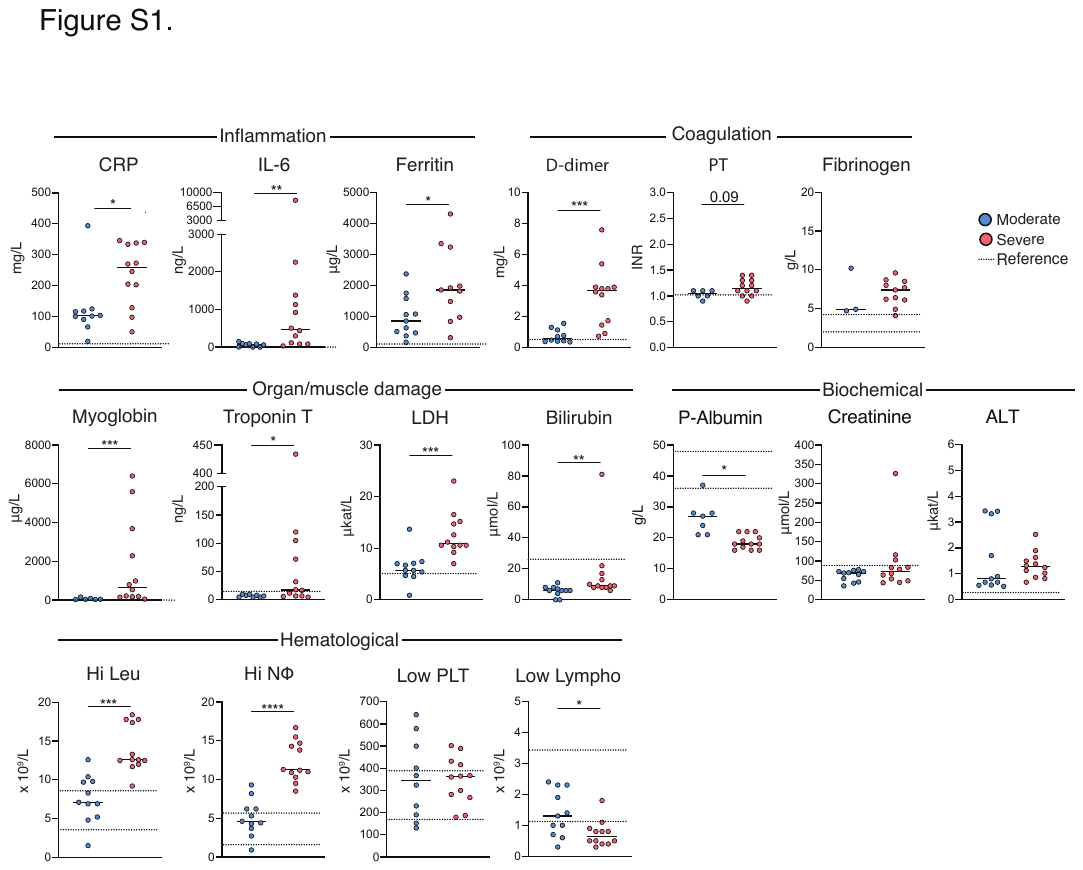

**Figure S1. Laboratory parameters in moderate and severe COVID-19 patients.** Serum levels of Inflammatory (CRP (mg/L), IL-6 (ng/L), Ferritin (μg/L)); Coagulation (D-dimer (mg/L), PT (INR), Fibrinogen (g/L)); Organ/muscle damage (myoglobin (μg/L), troponin t (ng/L), LDH (μkat/L), bilirubin (μmol/L)); Biochemical (P-Albumin (ng/L), creatinine (μmol/L), ALT (μkat/L)); Hematological (leukocyte (x10^9^/L), neutrophil (x10^9^/L), platelet (x10^9^/L), and lymphocyte (x10^9^/L) count)) markers in moderate and severe COVID-19 patients (moderate, n=3-11) (severe, n=11-12). CRP: highest C-reactive protein +/-24h from sampling (f.s.); IL-6: highest IL-6 +/- 5 days f.s.; Ferritin: highest ferritin +/-24h f.s.; D-dimer: highest D-dimer +/-24h f.s.; PT: highest prothrombin +/-24h f.s.; Fibrinogen: highest fibrinogen +/-24h f.s.; Myoglobin: highest myoglobin before sampling (b.s); Troponin T: highest troponin T +/-24h f.s.; LDH: highest lactate dehydrogenase b.s; Bilirubin: highest bilirubin +/-24h f.s.; P-Albumin: lowest P-albumin +/-24h f.s., Creatinine: highest creatinine +/-24h f.s.; ALT: highest Alanine Aminotransferase b.s.; Hi Leu: highest leukocyte count +/-24h f.s.; Hi NΦ: highest neutrophil count +/-24h f.s.; Low PLT: lowest platelet count +/-24h f.s.; Low Lympho: lowest lymphocyte count +/-24h f.s. Statistical differences were detected using Mann-Whitney test. Graphs are shown as median, *p < 0.05, ** p < 0.01, *** p < 0.001.

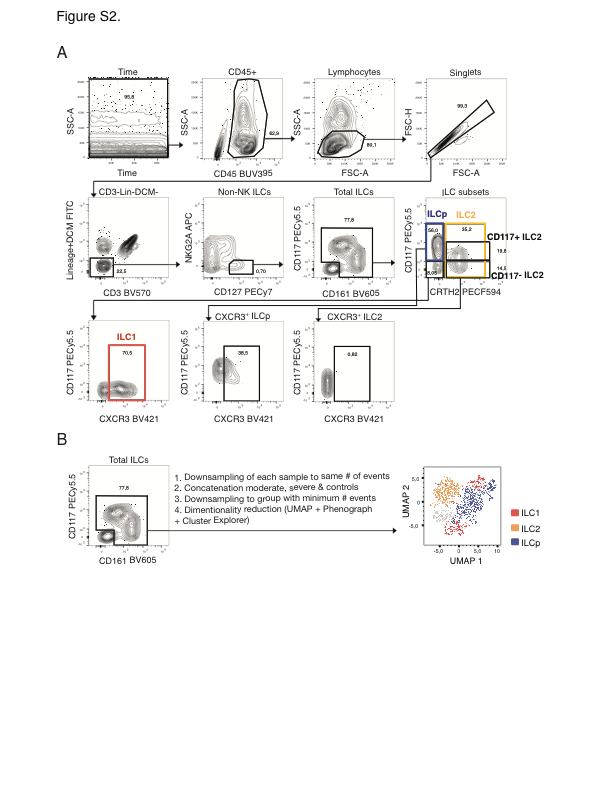

**Figure S2. Gating strategy for ILC identification in flow cytometry and UMAP analysis. (A)** Gating strategy used for the identification of ILCs as well as ILC1, ILC2 and ILCp subsets by flow cytometry. CXCR3 expression on ILCp and ILC2 (bottom row) serve as a positive and negative control, respectively, for the CXCR3 expression on ILC1 (left plot on bottom row). **(B)** Flow cytometry plot depicting the total ILC gate (left) that was used to perform the UMAP analysis (right).

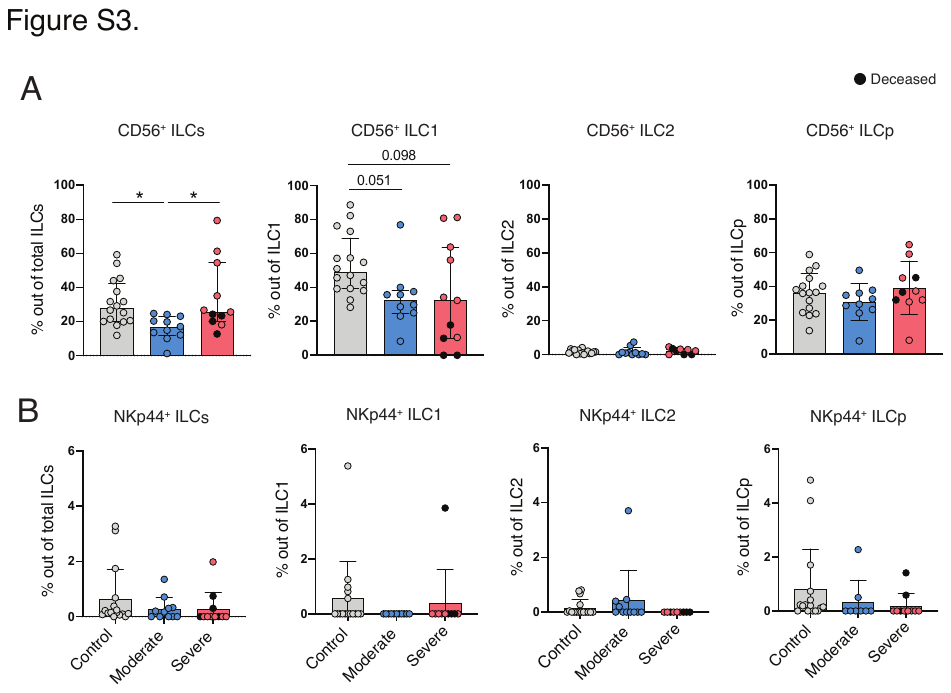

**Figure S3. CD56 and NKp44 expression on total ILCs and ILC subsets.** Bar plot summaries showing the percentages of **(A)** CD56 and **(B)** NKp44 in total ILCs and ILC subsets, ILC1, ILC2, and ILCp in control, moderate and severe COVID-19 patients. The number (n) of patients per subset is indicated in Fig. 3A-H. Statistical differences were tested using Kruskal-Wallis test followed by Dunn's multiple comparisons test. Bar graphs are shown as median ± IQR, *p < 0.05, ** p < 0.01, *** p < 0.001. Deceased patients in the severe group are indicated by a black dot.

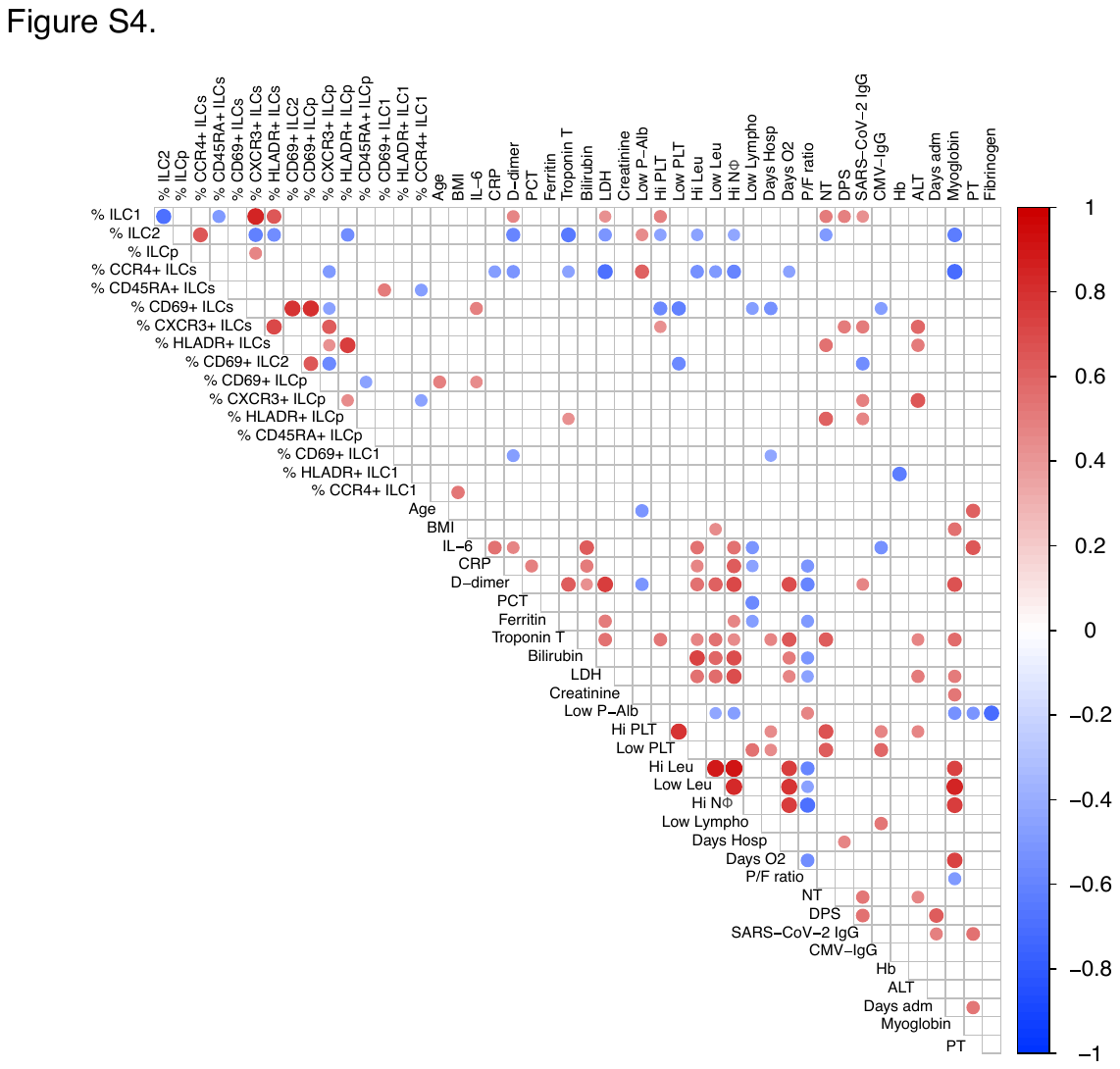

**Figure S4. Correlations of clinical and laboratory parameters with ILCs.** Spearman correlation matrix of clinical and laboratory parameters and ILC subsets in COVID-19 patients. The color of the circles indicates positive (red) and negative (blue) correlations that were statistically significant (p < 0.05) as measured by the Spearman’s rank correlation coefficient test. The color intensity and the size of the circle are proportional to the correlation coefficients. BMI: body mass index; IL-6: IL-6 levels at the time of sampling; CRP: highest C-reactive protein +/-24h from sampling (f.s.); D-dimer: highest D-dimer +/-24h f.s.; PCT: highest procalcitonin +/-24h f.s.; Ferritin: highest ferritin +/-24h f.s.; ; Troponin T: highest troponin T +/-24h f.s.; Bilirubin: highest bilirubin +/-24h f.s.; LDH: highest lactate dehydrogenase before sampling (b.s.); Creatinine: highest creatinine +/-24h f.s.; P-Albumin: lowest P-albumin +/-24h f.s.; Hi PLT: highest platelet count b.s.; Low PLT: lowest platelet count +/-24h f.s.; Hi Leu: highest leukocyte count +/-24h f.s.; Hi NΦ: highest neutrophil count +/-24h f.s.; Low Lympho: Lowest lymphocyte count +/-24h f.s.; Days Hosp: days of hospitalization until sampling; Days O2: days of oxygen treatment; P/F ratio: PaO2/FiO2 ratio; NT: titer of neutralizing antibodies; DPS: days post symptom debut to sampling; SARS-CoV-2 IgG: SARS-CoV-2 IgG antibodies; CVM-IgG: CVM-IgG IgG antibodies; Hb: lowest hemoglobin+/-24h f.s.; ALT: highest Alanine Aminotransferase b.s.; Days adm: days from symptom debut until admission in hospital; Myoglobin: highest myoglobin b.s.; PT: highest prothrombin +/-24h f.s.; Fibrinogen: highest fibrinogen +/-24h f.s.

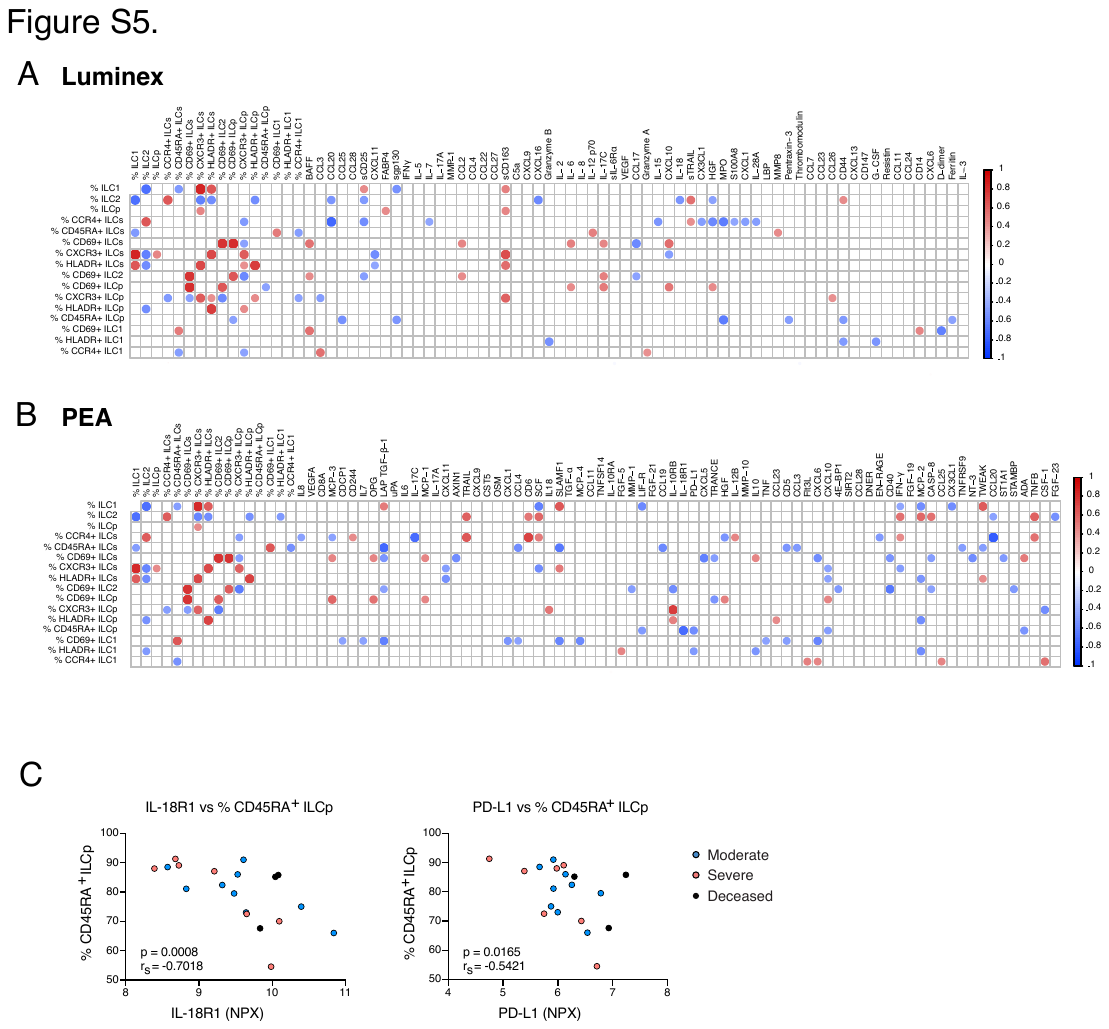

**Figure S5. Correlations of soluble factors with ILCs.** Spearman correlation matrix of the percentage of ILC subsets in COVID-19 patients and the level of soluble markers measured in serum by **(A)** multiplex immunoassays and **(B)** proximity extension assay. **(C)** Spearman’s rank correlations of serum IL-18R1 and PD-L1 relative levels, measured with proximity extension assay, and the percentage of CD45RA^+^ ILCp in COVID-19 patients. Data are shown as normalized protein expression (NPX). Blue circles: moderate COVID-19 patients (n=9); pink circles: severe COVID-19 patients (n=10); black circles: severe COVID-19 patients that passed away after sampling (n=4). p < 0.05 was considered statistically significant. r_s:_ Spearman's rank correlation coefficient. **(A-B)** The color of the circles indicates positive (red) and negative (blue) correlations that were statistically significant (p < 0.05) as measured by the Spearman’s rank correlation coefficient test. The color intensity and the size of the circle are proportional to the correlation coefficients.

**Table S1.** Antibody list used in flow cytometry

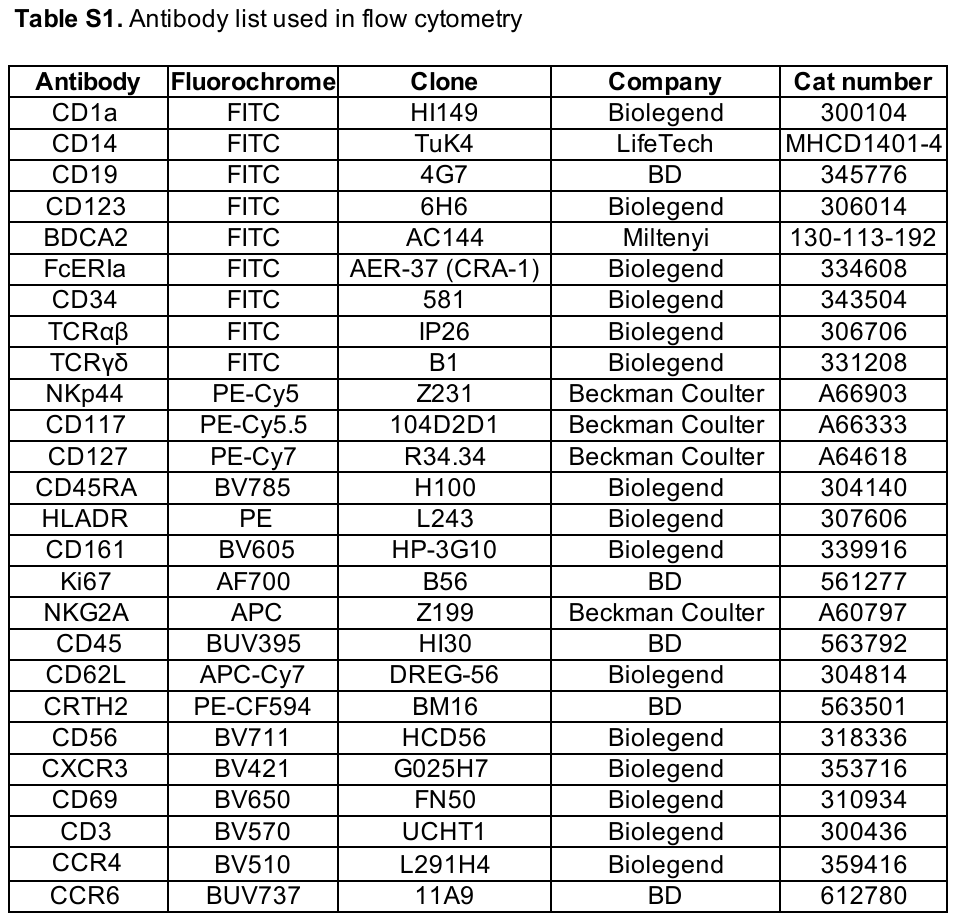
